# Genome-wide cell-free DNA termini in patients with cancer

**DOI:** 10.1101/2021.09.30.21264176

**Authors:** Norbert Moldovan, Ymke van der Pol, Tom van den Ende, Dries Boers, Sandra Verkuijlen, Aafke Creemers, Jip Ramaker, Trang Vu, Marieke F. Fransen, Michiel Pegtel, Idris Bahce, Hanneke van Laarhoven, Florent Mouliere

## Abstract

The structure, fragmentation pattern, length and terminal sequence of cell-free DNA (cfDNA) is under the influence of nucleases present in the blood. We hypothesized that differences in the diversity of bases at the end of cfDNA fragments can be leveraged on a genome-wide scale to enhance the sensitivity for detecting the presence of tumor signals in plasma. We surveyed the cfDNA termini in 572 plasma samples from 319 patients with 18 different cancer types using low-coverage whole genome sequencing. The fragment-end sequence and diversity were altered in all cancer types in comparison to 76 healthy controls. We converted the fragment end sequences into a quantitative metric and observed that this correlates with circulating tumor DNA tumor fraction (R = 0.58, p < 0.001, Spearman). Using these metrics, we were able to classify cancer samples from control at a low tumor content (AUROC of 91% at 1% tumor fraction) and shallow sequencing coverage (mean AUROC = 0.99 at >1M fragments). Combining fragment-end sequences and diversity using machine learning, we classified cancer from healthy controls (mean AUROC = 0.99, SD = 0.01). Using unsupervised clustering we showed that early-stage lung cancer can be classified from control or later stages based on fragment-end sequences. We observed that fragment-end sequences can be used for prognostication (hazard ratio: 0.49) and residual disease detection in resectable esophageal adenocarcinoma patients, moving fragmentomics toward a greater clinical implementation.

**One sentence summary:** cell-free DNA fragment end sequence analysis enhances cancer detection, monitoring and prognosis.

## INTRODUCTION

Liquid biopsies, and cell-free DNA (cfDNA) in particular, are actively investigated in clinical oncology. Genetic approaches, including screening for mutations and copy number aberration, are promising biomarker candidates for precision oncology *(1–4)*. Mutation-based detection of tumor-derived cfDNA is often hampered by technical and biological noise (the latter linked to the accumulation of mutations in normal cells) *(5, 6)*. For example, in elderly patients with TP53 mutant tumors, such as in esophageal adenocarcinoma (EAC), clonal hematopoiesis of indeterminate potential (CHIP) hampers the determination of the origin of cfDNA variants *(6, 7)*. In stage I-III patients with a low tumor fraction of cfDNA this requires a high sequencing depth for cfDNA, availability of buffy coat samples, tumor-informed sequencing or computational strategies to filter CHIP-derived variants *(8, 9)*. However, the complexity or costs of these methods are high and the clinical applicability is still limited. Methylation and fragmentomic sequencing have recently emerged as potentially sensitive and cost-effective alternatives *(10, 11)*.

During cell-death and mitosis, DNA can be cleaved at non-random locations and is subsequently released into the bloodstream *(12–15)*. This pool of cfDNA bears information about their cells of origin and mechanism of release *(16, 17)*. The type of DNase cleaving the DNA is dependent on the nucleosome organization, and the presence or absence of cofactors, resulting in distinct fragment sizes and Fragment End Sequences (FrESs) *(12, 13)*. The size distribution of cfDNA, with a mode of ∼167 bp and multiples thereof, is related to the wrapping of DNA around the nucleosomes *(18)*. An increase in the proportion of shorter fragment sizes (< 150bp) can be observed in the presence of tumor, which may help monitor or forecast disease outcome *(19–21)*. Furthermore, a genome-wide analysis of the size profile can identify cancer from different types and stages *(22, 23)*, which could complement methylation or nucleosome footprinting analysis of cfDNA *(24, 25)*. Studies of fragment-end sequence profiles revealed the predominance of C-rich 5’ end motifs, linked to the activity of DNASE1L3 in apoptotic cells and in plasma *(12)*. The proportion of fragments ending with a C-rich motif is decreased in patients with cancer, and they show a higher sequence diversity in their fragment ends *(26)*. Information on the diversity and clinical utility of FrESs in oncology remain limited *(26)*. Open and reproducible workflows to investigate FrESs are also missing.

We aim to improve the sensitivity of cfDNA-based non-invasive cancer analysis by leveraging the biological differences that determine the FrES. We hypothesized that the FrESs patterns correlate with the ctDNA tumor fraction in plasma, and can be utilized for the detection, prognostication and monitoring of cancer. To test this, we established a genome-wide catalogue of cfDNA fragment end sequence patterns of a large cohort of cancer patients with 18 different cancer types. Using these biological patterns, we developed the Fragment End Integrated Analysis (FrEIA) score to quantitatively evaluate the tumor fraction in liquid biopsy samples using low coverage whole-genome sequencing (WGS). We showed that the fragment-end sequences of cfDNA can be leveraged to enhance the detection and monitoring of cancer. We demonstrated the prognostic value of the FrEIA score in patients with resectable EAC at baseline and after surgery. Finally, by integrating multiple FrES with and without machine learning, we demonstrate that FrES can be used to classify cancer types and stages.

## RESULTS

### Surveying the fragment-ends features of tumor cfDNA

We generated a catalogue of cfDNA fragmentomic features in 572 plasma samples from 319 patients with 18 different cancer types, an additional 76 control samples and 10 samples from patients with lung nodules not otherwise classified **(Table S1)**. 313 samples were acquired at baseline prior to any treatment, while 259 were collected after various lines of treatment. Sequencing data for 243 of the samples were retrieved from a previous study *(22)* (designated **R samples**), while 416 are newly collected from two studies **(Table S2;** the esophageal cancer samples are designated **E**, and the cohort containing mostly lung cancer cases **L samples)**.

To characterize and assess the FrES patterns of cfDNA from genome-wide sequencing we developed the FrEIA toolkit (see Methods). cfDNA fragments from all three datasets were categorized based on the frequency of either their 5’ nucleotide (4 features), their 5’ and 3’ nucleotide (4×4 = 16 features) and the first three bases on the 5’ end (4^3^ = 64 features) (**Figure 1A**). Datasets with both control and cancer samples (R and L) were used to characterize the FrES differences between patients with cancer and control individuals. The most abundant 5’ fragment ending nucleotide for both cancer patients and control individuals was C (cancer patients: median 33% of fragments, control subjects: median 36% of fragments) followed by T (cancer patients median 30%, control subjects median 29% of fragments) and A (cancer patients median 18%, control subjects median 17% of fragments) (**Figure S1A**). In the cancer samples we detected a significant decrease in the proportion of fragments starting with C (log_10_(cancer/control) = -0.03, p = 3.92*10^-12^, two-sided Mann-Whitney U test) and an increase of fragments starting with A (log_10_(cancer/control) = 0.02, p = 1.87*10^-8^, two-sided Mann-Whitney U test) and T (log_10_(cancer/control) = 0.01, p = 0.008, two-sided Mann-Whitney U test) (**Table S3)**. Fragments with a 5’ C or a 3’ G end (the latter being a C on the Crick strand) were significantly decreased in cancer (p < 0.05). Any other combination was increased in cancer, except the 5’ C - 3’ C, the 5’ G - 3’ C, the 5’ G - 3’ G and the 5’ T - 3’ A combinations, which exhibited no significant difference between cancer and control (**Figure S1B, Table S4**). Twenty seven of the 5’ trinucleotide sequences decreased significantly in cancer samples compared to control, while twenty increased (alpha = 0.01; **Figure 1B, Table S5**).

**Figure 1.**
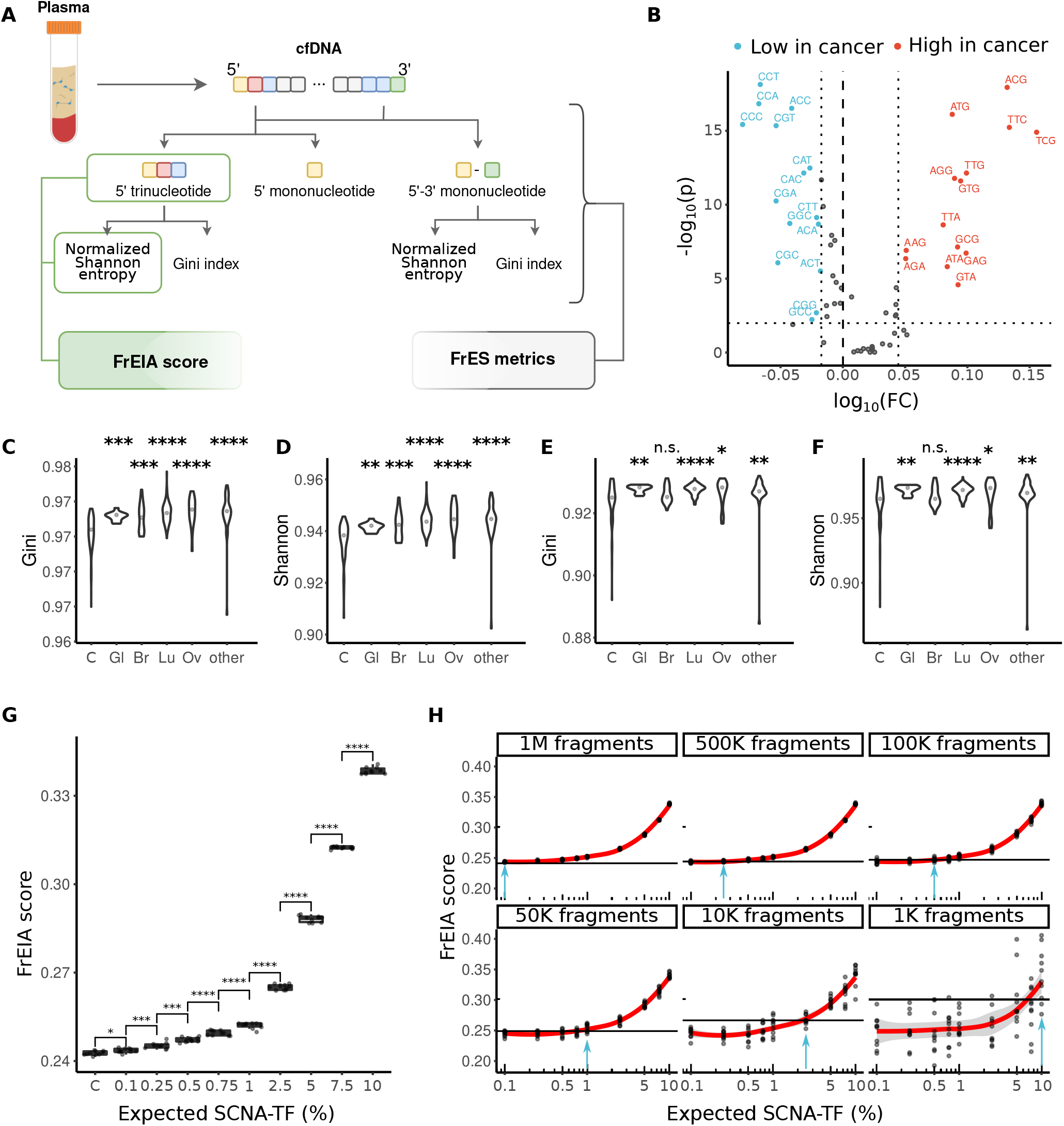
cfDNA fragments termini composition and diversity are altered in cancer. **A**. Schematic of the FrEIA pipeline. The FrEIA score is a metric derived from the 5’ trinucleotides and the normalized Shannon entropy. The FrES metrics include the diversity and the proportion of fragment end sequences (see Methods). **B**. The fold change of trinucleotide FrESs in baseline cancer compared to control’s trinucleotide FrESs that pass the significance threshold of 0.01 calculated by the two-sided Mann-Whitney U test (horizontal dotted line) and their fold change is lower than the 25^th^ percentile (left vertical dotted line) are in blue while those for which the threshold is higher than 75^th^ percentile (right vertical dotted line) are in red. The median diversity in (**C**. and **D**.) 5’ trinucleotide and (**E**. and **F**.) 5’-3’ mononucleotide of cfDNA from cancer samples of the R sample set is increased compared to controls. Cancer types with less than 10 samples at baseline were merged into the “other” category (Bonferroni corrected p-values. *: p < 0.05, **: p < 0.01, ***: p < 0.005, ****: p < 0.0001). Sample types: C: control, Gl: glioblastoma, Br: breast cancer, Lu: lung cancer, Ov: ovarian cancer. **G**. At 1M fragments (of the *in silico* dilutions) the FrEIA score increases with the TF. Samples with as low as 0.1% TF are detectable, with a significantly higher FrEIA score than controls (p-values. *: p < 0.05, ***: p < 0.005, ****: p < 0.0001). **H**. The change of detectable TFs by the decrease of sequencing depth in the *in-silico* dilutions. The horizontal line represents the detection threshold, which is calculated as the mean + 2*SD of the controls at the specific depth. Blue arrows indicate the TF with a mean intersecting the threshold: 1M fragments: 0.241, 500k fragments: 0.244, 100k fragments: 0.247, 50k fragments: 0.248, 10k fragments: 0.266, 1k fragments: 0.3.

To quantify with a single metric the extent of diversity of cfDNA fragment-ends, we calculated a normalized Shannon entropy and a Gini index of the 5’ trinucleotide and 5’-3’ mononucleotide sequences (see Methods). We detected an increase in median diversity of FrES for cancer patients in comparison to control individuals (*normalized Shannon entropy:* 5’ trinucleotide: 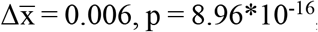, 5’-3’ mononucleotide: 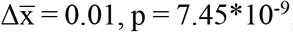; *Gini index:* 5’ trinucleotide: 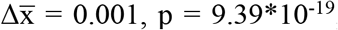, 5’-3’ mononucleotide: 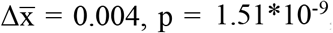, two-sided Mann-Whitney U tests). This increase was observed for each cancer type analyzed (**Figures 1C-F**).

As plasma cfDNA from cancer patients exhibited altered trinucleotide FrES proportions and diversity (from now on FrES metrics) in comparison to healthy individuals, we integrated these metrics into a single quantitative measurement called the *FrEIA score* (**Figure 1A**). The FrEIA score was calculated by combining the normalized Shannon entropy of samples with the proportions of 29 selected trinucleotides FrES. The trinucleotides were selected if they had a fold change compared to control above (log_10_(FC) > 0.044) or below (log_10_(FC) < -0.017), and if the change was significant (p < 0.001). For this calculation, only baseline cancer samples with a TF higher than 10% (n_cancer_ = 56, n_control_ = 76) were used (**Figure 1B**) (see Methods). To determine the detection limits of the FrEIA score, we created a series of 10 *in silico* dilutions with known SCNA-TF (see Methods) 10 iterations each. The FrEIA score of samples with an expected SCNA-TF of 0.1% (and above) were significantly higher than the FrEIA score of control samples using 1M fragments (p < 0.05, two-sided Mann-Whitney U test) (**Figure 1G**). In addition, we evaluated if the FrEIA score could be used with reduced coverage of sequencing, and downsampled our sequencing data from 1 M cfDNA fragments down to 100k, 10k and 1k fragments. The FrEIA score was able to distinguish TF higher than 0.5% at 100k, TF higher than 2.5% at 10k and TF higher than 10% using only 1k cfDNA fragments (**Figure 1H**). Based on these results we conclude that the FrEIA score can be a sensitive and potentially cost-effective addition to other analysis performed using low coverage WGS.

### The FrES is altered by the ctDNA tumor fraction in plasma and by the cancer type

The median FrEIA score of 313 cancer samples from different types collected pre-treatment is increased compared to 76 control samples (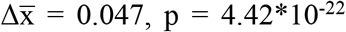, two-sided Mann-Whitney U test). A similar increase was detected for cancer types with more than 10 samples at baseline (n = 5 cancer types) (**Figure 2A**).

**Figure 2.**
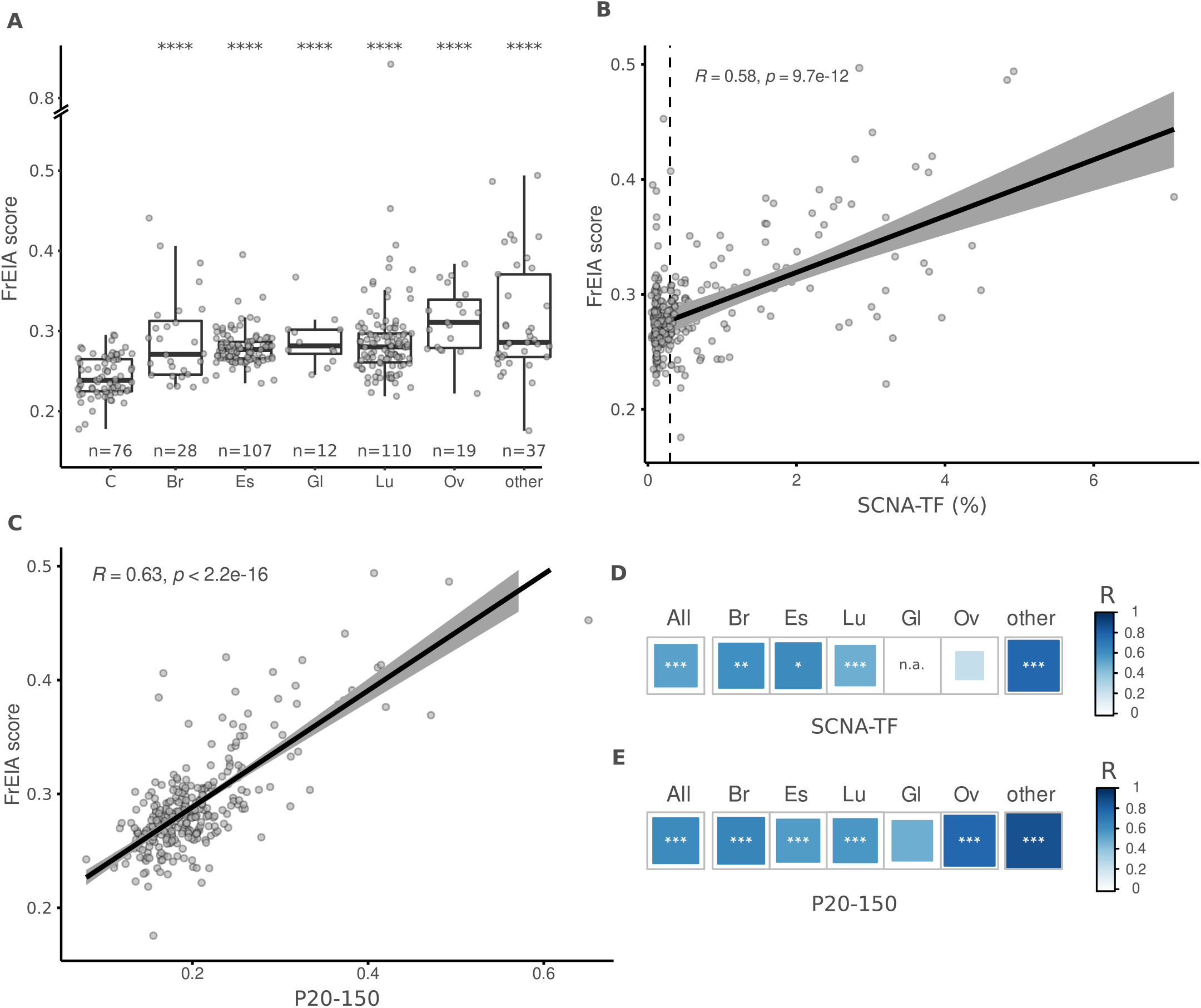
FrEIA score is increased in cancer and differs by cancer type. **A**. Mean FrEIA score of baseline cancer samples is significantly increased (Bonferroni corrected p-values. ***: p < 0.005, ****: p < 0.0001) in every cancer type compared to controls. **B**. Spearman correlation coefficients of FrEIA scores with SCNA-TF and **C**. with proportion of fragments between the size range of 20-150 (P20-150) for every baseline cancer sample. **D**. Spearman correlation coefficients of FrEIA scores with SCNA-TF and **E**. with proportion of fragments between the size range of 20-150 (P20-150) by cancer type for samples collected at baseline. For SCNA-TF the correlation is calculated only for samples with a TF >= 3%. Abbreviation of cancer types: C: control; Br: breast; Gl: glioma; Lu: lung; Ov: ovarian. P-values: *: p < 0.05, **: p < 0.01, ***: p < 0.005.

The FrES metrics and the FrEIA score were compared to the ctDNA tumor fraction (TF) to evaluate their potential as quantitative markers. We calculated the ctDNA TF represented by either the SCNA-TF or the proportion of fragments between 20 and 150 bp (P20-150) using the same sequencing data. The proportion of fragments starting with TTC showed a strong positive correlation (Spearman R ≥ 0.6, p < 0.001) with both measures of TF, the proportion of fragments starting with ATG showed a strong positive correlation with P20_150 and a moderate correlation with SCNA-TF (Spearman R between 0.4 - 0.6, p < 0.001) while ACG and TCG showed a strong positive correlation with SCNA-TF and a moderate correlation with the P20_150 **(Table S6 and Table S7**). These observations remained consistent irrespective of the cancer type **(Figure S2, Table S8 and Table S9)**, suggesting a common mechanism of cfDNA cleavage in cancer. Moreover, the FrEIA score was positively correlated to the TF metrics (SCNA-TF: Spearman R = 0.58; P20-150: Spearman R = 0.63; p < 0.01) (respectively, **Figure 2B** and **2C**), which could be observed for all cancer types with the exception of gliomas (**Figure 2D, 2E and Figure S2)**. Based on this we conclude that the FrEIA score can be used as a quantitative marker of the presence of tumor-derived DNA in the plasma of patients with various cancer types.

### The FrES and the FrEIA score enables detection of cancer stage

Next, we evaluated whether the FrEIA score could be used for the early detection of cancer. We selected a subset of our cohort with lung cancer, where all cases were collected from the same clinical center under the same pre-analytical conditions and methods. This sub-cohort was composed of samples from patients with lung nodules (or other lesions) (n = 10), patients with early-stage lung cancer (stages I and II, n = 14), stage III (n = 27) and stage IV of the disease (n = 66) (**Figure 3A**). All cancer samples, irrespective of their stage, revealed a significant mean increase in FrEIA score compared to either the samples from control subjects or patients with nodules (stages I-II - nodule: 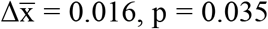; stage III - nodule: 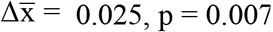; stage IV - nodule: 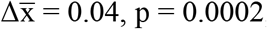, two-sided Mann-Whitney U tests) (**Figure 3A**). The FrEIA scores for samples of stage IV patients were also higher than for the samples of stage I-II patients (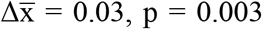, two-sided Mann-Whitney U test). We could not detect significant variation in the FrEIA scores of nodule samples compared to controls (p = 0.36, two-sided Mann-Whitney U test). The higher analytical sensitivity of the FrEIA score compared to that of SCNA-TF detection, results in a better separation between stages compared to the SCNA-TF based method (**Figure 3B)**, even at a TF below the 3% threshold of SCNA-TF detection **(Figure S3A**). This suggests that a FrEIA score-based classification of cancer stages can remain sensitive even at low TF, even if this needs further investigation in a larger cohort of samples.

**Figure 3.**
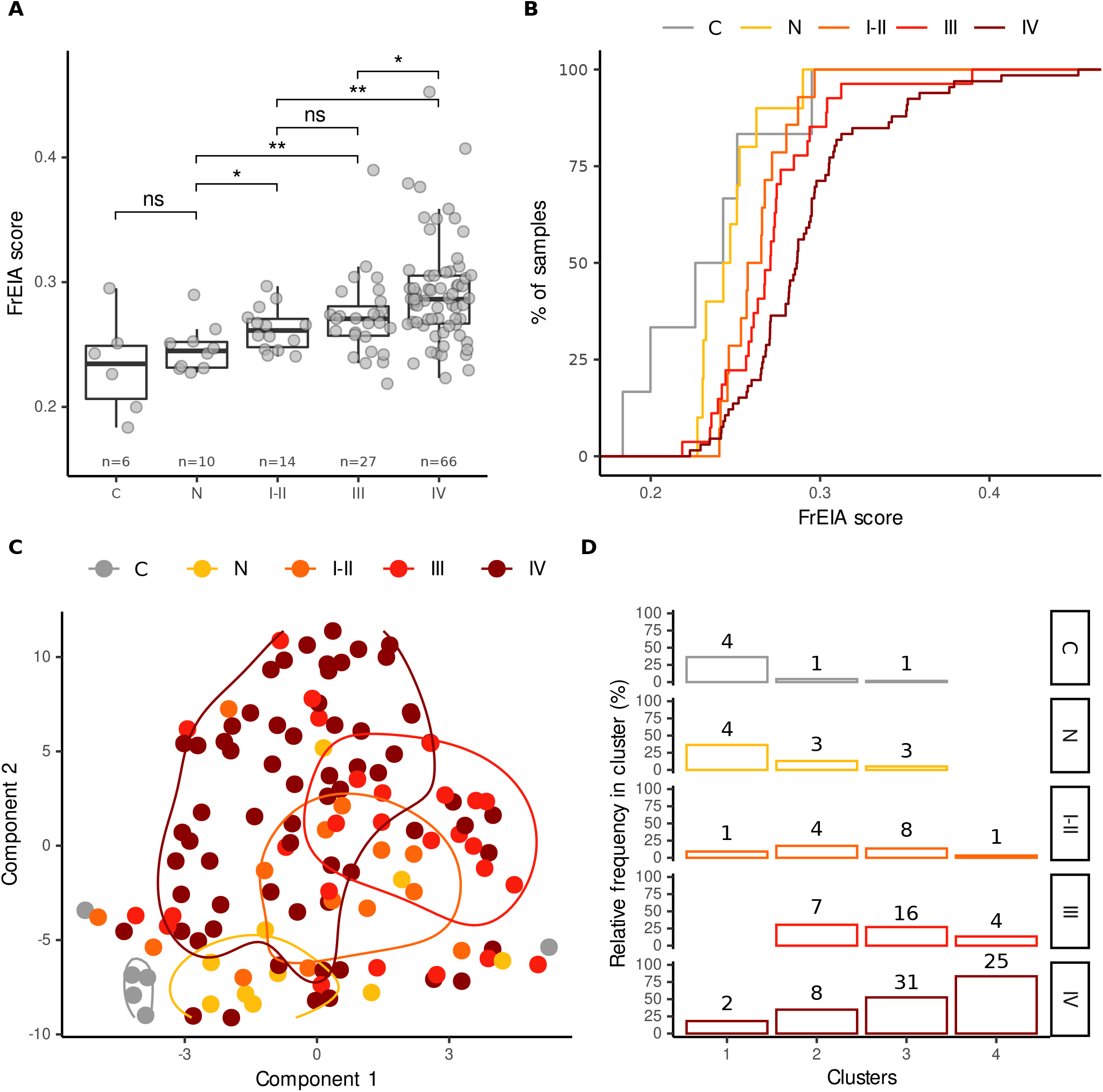
FrEIA score and FrES are altered in early- and late-stage lung cancers. **A**. FrEIA score of controls (C), lung nodules (N) and lung cancer stages (I-II, III and IV) compared. Bonferroni corrected p-values. n.s.: not significant, *: p < 0.05, **: p < 0.01. **B**. FrEIA score distinguishes cancer stages from controls and nodules. **C**. t-distributed stochastic neighbor embedding (t-SNE) plot, generated using FrES metrics for the cohort of 106 lung cancer samples with associated density contours. **D**. k-means clustering on the same data set and features assuming 4 clusters. Numbers on top of the bars represent sample count. Cancer stages: C: control (n = 6), N: nodule or lesion (n = 10), stages I-II: n = 14, stage III: n = 27, stage IV: n = 66.

To assess the potential of FrES metrics and FrEIA score for detecting early-stage cancer we used unsupervised clustering (t-SNE) on the samples from the lung cancer cohort (**Figure 3C**). The L cohort also contained a small number of controls (healthy n = 6, nodules n = 10) all recruited in the same center and processed using the same pre-analytical methods to reduce biases resulting from batch effects (**Figure S3B**). We used a k-means algorithm to cluster the L samples in four clusters and observed that 73% of the first cluster (n = 8) is from non-cancer samples (healthy controls and nodules cases). Cluster 2 to 4 accounted for 106 samples, with among them were 98 cancer cases (92%). 93% (n=13) of the early-stage (I-II) cases clustered together with other cancer stages. The proportions of stages in each cluster showed a similar trend, where most of the control samples and nodules clustered together, while the proportion of cancer samples gradually increased in the clusters (**Figure 3D, Figure S3C**).

### Prognostic potential of FrES and FrEIA score

To evaluate the prognostic value of FrEIA in a clinical setting, we tested cfDNA from resectable EAC patients, where serial ctDNA detection has been shown to predict adverse outcome *(7)*. Here we assessed the potential of the FrEIA score in 293 resectable EAC samples from 2 clinical cohorts: a neoadjuvant chemoradiotherapy (nCRT) cohort (n=70 patients, n = 149 plasma samples) receiving standard of care carboplatin combined with paclitaxel based nCRT and a cohort of patients who participated in the phase II PERFECT trial (n=40 patients, n = 144 samples, see Methods) received nCRT in combination with a PD-L1 inhibitor *(27)*. Both cohorts included EAC stage II (n=125 samples) and stage III (n=168 samples). Plasma samples were collected longitudinally before and after chemoradiation, and also postoperatively in the PERFECT cohort (**Figure 4A**). We removed one patient with an atypical early progression (93 days from the start of the study, median progression time 589 days). Using sWGS, the TF and other cfDNA patterns can be monitored and compared for each timepoint (**Figure 4B** and **Figure S4**).

**Figure 4.**
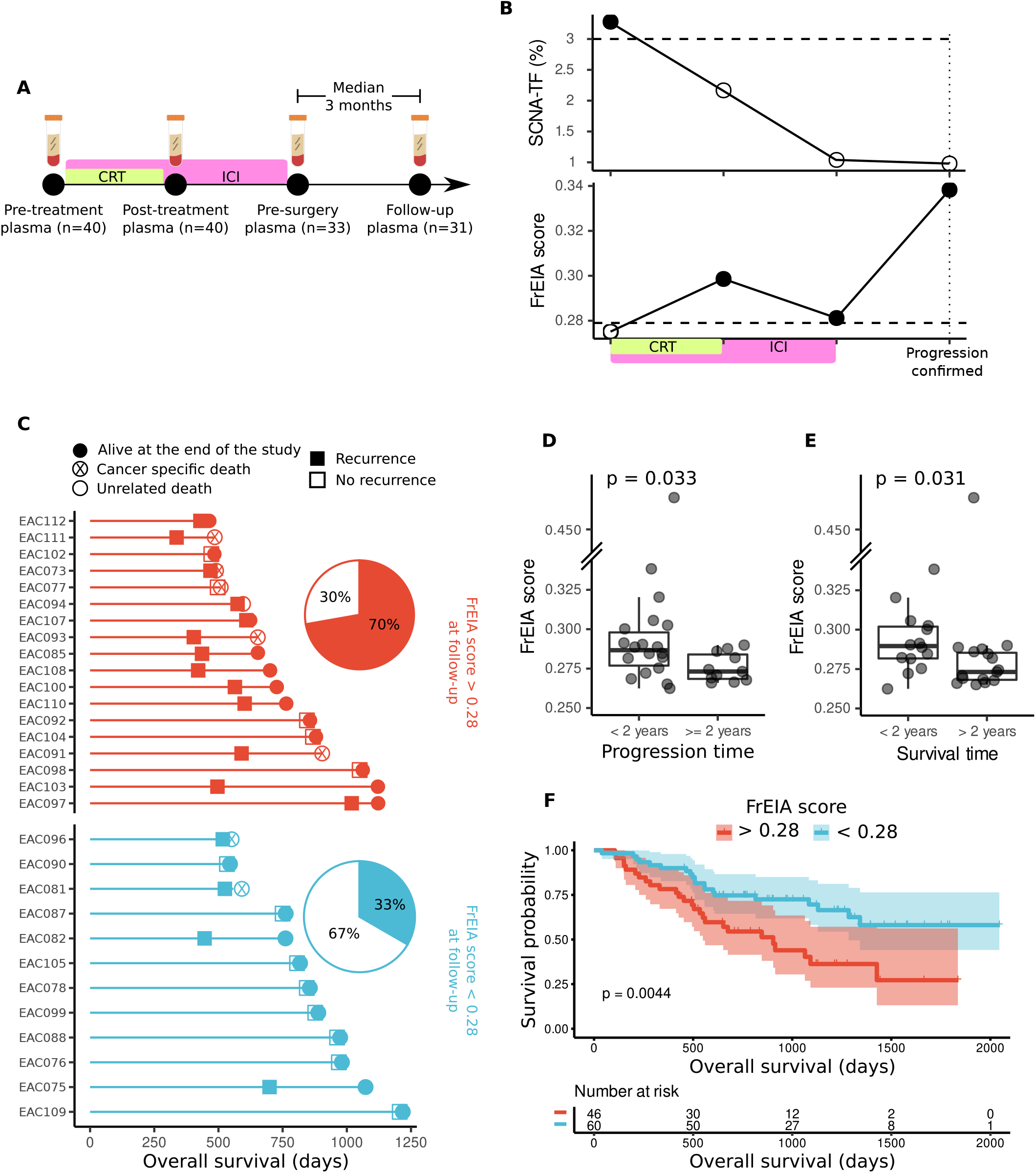
The FrEIA score is higher in esophageal cancer patients with early progression after surgery and is prognostic for survival at baseline. **A**. Schematic of the clinical timeline of the samples from the PERFECT cohort (n = 40 patients, as part of the C, D and E cohort). Plasma samples were collected before and following chemoradiotherapy (CRT) in combination with immune checkpoint inhibition therapy (ICI) and preceding surgery. Follow-up plasma samples were collected at a median of 3 months after surgery. **B**. The change of SCNA-TF and of the FrEIA score of a single patient during the clinical timeline. Horizontal dashed lines represent the detection thresholds for the SCNA-TF (3% TF) and the FrEIA score (0.28). Filled circles are values above, while empty circles are values below the thresholds of detection. **C**. Event chart showing progression detection times based on CT-scans and survival of the patients with a FrEIA score above and below the 0.28 threshold detected at follow-up. The pie-charts show the proportions of patients with (filled slices) and without (empty slices) progression in the two groups. **D**. The FrEIA score of patients with recurrence earlier and later than two years following the start of the study. FrEIA score measured at follow-up. **E**. The FrEIA score of patients with a survival shorter and longer than two years following the start of the study. FrEIA score measured at follow-up. **F**. Kaplan-Meier survival curves showing overal survival and risk assessment of the nCRT and PERFECT cohorts with the FrEIA score below and above the detection threshold detected pre-treatment.

The detection of residual disease can help select patients who may benefit from adjuvant treatment. However, detection of residual disease using liquid biopsies is particularly challenging shortly after surgery, and often requires complex approaches such as tumor-guided sequencing *(28, 29)*. But it could be informative for the application of neoadjuvant chemotherapy in patients with high risk of progression. We calculated the FrEIA score of samples collected ∼3 months after surgery, and split the samples in two groups with the FrEIA score lower (n = 12) and higher (n = 18) than 0.28, a stringent threshold selected based on the synthetic dilution series. Thirteen of the eighteen patients with a high FrEIA score (>0.28) had progression following sampling, while in those with a lower score (≤0.28), only four out of twelve patients showed progression (**Figure 4C**). Furthermore, patients with early progression or shorter survival had a significantly higher FrEIA score than those with later progression or longer survival (respectively, mean FrEIA score = 0.29, SD = 0.008 compared to 0.27 SD = 0.04, p = 0.033 and mean FrEIA score = 0.29, SD = 0.019 compared to 0.28 SD = 0.047, p = 0.031 respectively) **(Figure 4D, Figure S5 and Figure 4E**, respectively).

To test the prognostic value of the FrEIA score we split our pre-treatment samples from both cohorts in two groups with the FrEIA score lower (n = 60) and higher (n = 46) than the threshold set at 0.28. Patients with a FrEIA score above this threshold showed a median survival of 904 days, while the number of patients with a FrEIA score below the threshold have a reduced risk of dying from cancer (hazard ratio: 0.49, p = 0.004, 95% CI [0.23, 0.75]) (**Figure 4F**). A high baseline FrEIA score showed similar predictive power for patient survival, recurrence or incomplete response (**Figure S6**), both of which are determined later during the clinical timeline, suggesting that the baseline FrEIA score has prognostic potential for survival.

### Cancer and cancer type classification based on FrES metrics and FrEIA score

The primary use of cfDNA fragmentation features in oncology was for improving the detection of cancer. Fragment end sequences were used to classify cancer from healthy controls in a cohort of hepatocellular carcinoma cases *(26)*. We developed a support-vector machine-based (SVM) learning algorithm for cancer classification using either the FrES metrics or the FrEIA score (**Figure 5A**). FrES metrics with the best predictive value were first tested and selected for further classification steps (**Figure S7**.). Baseline samples from the three datasets (cancer samples n = 313, controls n = 76) were randomly split (80% for training/validation and 20% for testing) (see Methods). On the training/validation set, 10-fold cross validation was carried out to perform hyperparameter optimization. The final models were trained using the optimized hyperparameters and the performance of the models were retrieved from testing the unseen test set. The model using the FrES metrics as input features performs better when classifying pre-treatment cancer samples from control samples (mean AUROC = 0.99, SD = 0.01), having a mean positive predictive value (PPV) of 99% (SD = 0.008) and a mean negative predictive value (NPV) of 96% (SD = 0.04) over 10 iterations of testing (**Figure 5B and 5C**). While the model using the FrEIA score showed a lower mean PPV (86%, SD = 2%), and mean NPV (87%, SD = 13%) (**Figure S8**).

**Figure 5.**
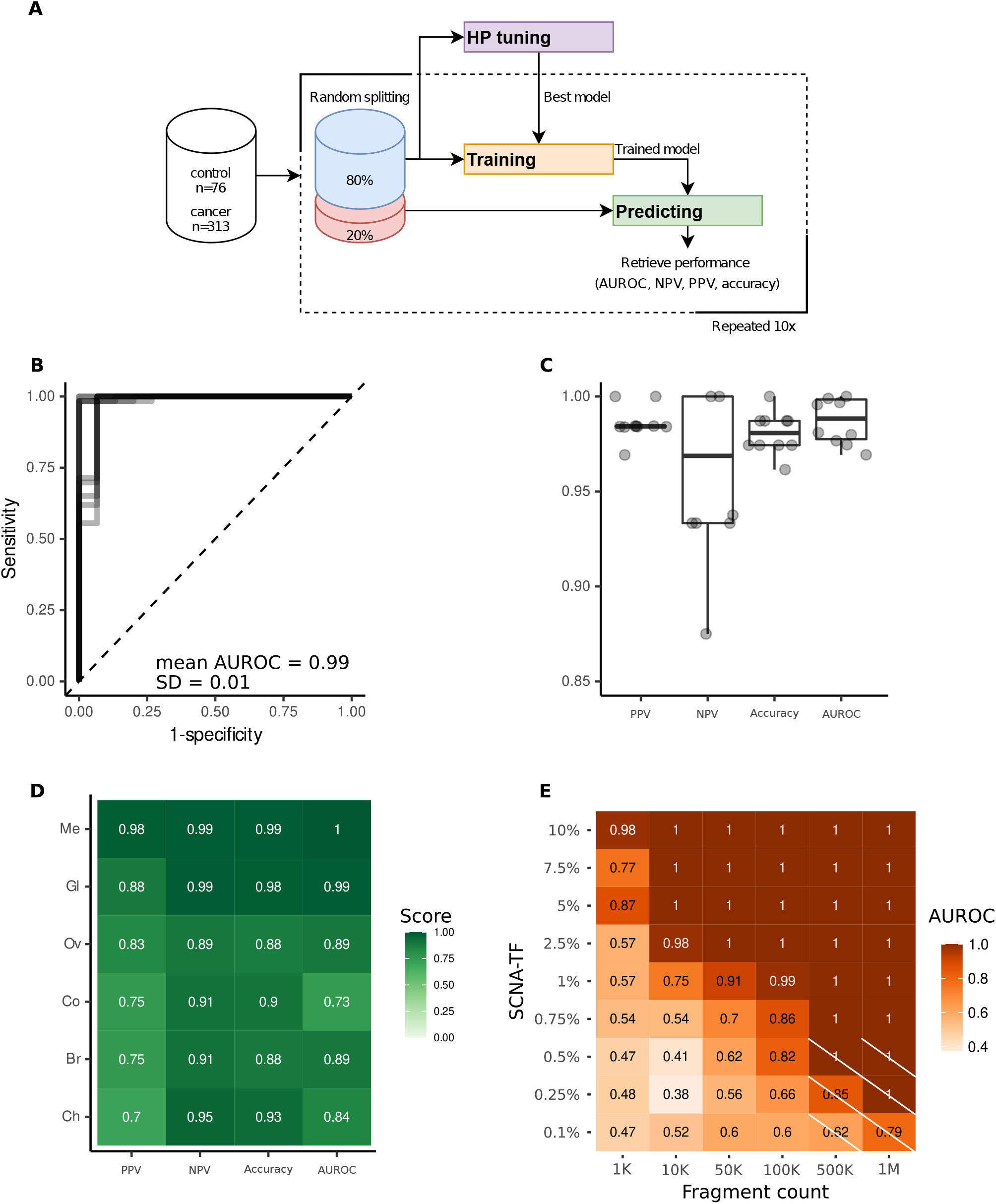
FrES metrics and the FrEIA score classify cancer types and stages. **A**. Workflow used for training and predicting Support Vector Machine (SVM) based models using 10-fold cross-validation with random sampling on the samples collected at baseline (n = 313). **B**. Receiver Operating Characteristic (ROC) curve from the 10 iterations of the trained cancer (n = 313) versus control (n = 76) model. **C**. Performance metrics of the model classifying cancer from control on 10 iterations of testing. PPV: positive predictive value, NPV: negative predictive value, AUROC: area under the receiver operating characteristic curve. **D**. Performance of the FrES-metrics-based model classifying cancer types of the R dataset. Me: melanoma, Ov: ovarian cancer, Gl: glioblastoma, Br: breast cancer, Ch: cholangiocarcinoma, Co: colorectal cancer. **E**. Detection limits of the SVM model tested on an *in-silico* generated dilution series with controlled TF and increasing sequencing depths. Crossed-out cells hold values where the number of true-positives and false-positives were 0 and thus a PPV could not be computed.

To evaluate if a model can classify one cancer type from all the others (and not healthy controls), we selected 5 types (breast, cholangeal, colorectal, ovarian cancer, glioma and melanoma) that were represented by more than 10 samples in the full dataset (pre- and post-treatment). To eliminate biases caused by pre-analytical batch effects (see **Supplementary Methods** and **Figure S3A**) we only used the R dataset for this classification, for which we detected no inter-type classification bias using a t-SNE clustering **(Figure S9)**. The model using the selected FrES metrics (**Figure S10**) performed the best on the melanoma (mean PPV = 98%, SD = 6%; mean NPV = 99%, SD = 3% on 10 iterations of testing) and glioblastoma cancer cases (mean PPV = 88%, SD = 19%; mean NPV = 98%, SD = 2% on 10 iterations of testing) when classifying these from the other cancer types (**Figure 5D and Figure S11)**.

To determine the detection limits of the FrEIA score-based and FrES-metrics-based classifiers, we evaluated the SVM models using *in-silico* dilutions of one cancer sample in one control sample. The FrEIA score-based model performed the best (**Figure S12**), with a mean PPV and NPV of 90%, and a mean AUROC of 91% at 1% TF and 100k fragment. The performance of cancer detection improves as the number of cfDNA fragments or the tumor fraction increases (**Figure 5E**). Using the FrEIA score with a SVM model, a 3x improvement can be achieved over the detection limit of ichorCNA *(30)* and a 4x improvement compared to the fragment-end diversity alone *(26)*.

## DISCUSSION

Using a pan-cancer dataset (18 cancer types, 572 plasma samples) and low coverage WGS, we demonstrate that the non-random cleavage of cfDNA fragment-ends is a common phenomenon in the plasma of cancer patients, irrespective of their cancer type and can predict patient outcome.

Our new pipeline, called FrEIA, allows the recovery of cfDNA FrES from genome-wide sequencing data in a reproducible way. FrEIA can be used on low coverage WGS, a cheap and highly standardized method for cfDNA analysis *(22, 30, 31)*, as well as higher depth WGS *(23, 24, 32)*. We identified biological features that could determine the presence and amount of ctDNA in plasma samples, without a priori knowledge of somatic aberrations and using low coverage WGS. The diversity of the FrES in cancer was increased compared to controls, which was persistent for each cancer type, and irrespective of the method used to calculate the fragment-end diversity. Based on these observations, we developed the FrEIA score, and showed that it is increased for cancer samples, for each cancer type and stage, and that it correlates with the ctDNA tumor fraction.

Using *in-silico* dilution series, from 1M fragments we determined that FrEIA can discriminate tumor samples from healthy controls down to ∼0.25% tumor fraction, in comparison to ∼3% tumor fraction for low coverage WGS methods based on SCNA analysis alone *(30)*. Therefore, our method can be used to detect the tumor signal for cancers lacking or with undetectable copy number aberrations, and also early-stage cancer (I-II) with low tumor fraction. FrEIA also achieves high sensitivity for detecting subtle changes in FrES from tumor signal using minute amounts of cfDNA fragments, and achieves an analytical sensitivity of 1% TF from only 50k fragments.

Our work provides strong evidence that the composition in bases at the end of cfDNA fragments in the plasma of multiple cancer types is altered in comparison to healthy controls. This difference was previously observed in a cohort of hepatocellular carcinoma *(26)*. Using FrEIA on 572 plasma samples from 18 different cancer types, we identified that fragment-end sequences are different in cancer compared to healthy controls on both the mononucleotide and the trinucleotide levels. ctDNA tumor signal is correlated to specific composition of FrES in the plasma of cancer patients. In plasma from high ctDNA tumor fraction, cfDNA fragments ending in A and T are significantly overrepresented in comparison to low ctDNA samples. Technical and pre-analytical conditions pose potential limitations to FrES usage, and could generate false conclusions arising from batch effects if not carefully examined. Similarly, the choice of library preparation with either single-stranded and double-stranded DNA *(33)*, or PCR and PCR-free *(34)* could impact the size distribution of cfDNA, with an anticipated method-dependent bias in the FrES composition. Furthermore, FrES analysis represents an indirect detection of tumor signal, and can be obscured by other clinical conditions that affect cfDNA release in the bloodstream *(35–37)*. Another limitation is the number of nucleotides chosen for analysis, as the stretch of DNA carrying tumor-specific signal is currently not clearly defined. Improving the characterization of the fragment-ends will be important, especially for future work combining analysis of ctDNA with that of other blood components such as extracellular vesicles, proteins and tumor-educated platelets.

The FrEIA pipeline extracts 84 cfDNA-derived metrics from cfDNA sequencing data that can be leveraged using machine learning classifiers to improve the detection of cancer in challenging clinical scenarios. These metrics combined using a SVM classifier on a cohort of 648 samples (572 cancer and 76 control samples) lead to a mean AUROC = 0.99 (SD = 0.01). These FrES metrics can be used alone, or in combination with other cfDNA features generated during the same run of sequencing (e.g., SCNA, fragment size distribution, genome-wide fragmentation, transcription factor binding sites, structural and topological analysis) *(10, 35, 38)*.

At the difference of methods relying on supervised machine learning to leverage cfDNA features, and thus prone to the inherent biases of this type of classification, here we demonstrated that FrEIA enables unsupervised classification of early and late-stage cancers. In a cohort of 106 lung cancer cases, all from the same clinical center, including 14 early stage samples we could identify groups consisting of mainly cancer cases (one group with 30/30 cancer cases and 0 controls, one group with 55/59 cancer cases three nodule and one control sample, one group with 19/23 cancer cases, tree nodule and one control cases), while controls and samples from patients with nodules aggregated together in a separate cluster.(one group of 4/11 control and 4/11 nodule and 3/11 cancer cases).

Beyond cancer classification, the FrES metrics recovered using FrEIA can be used in realistic clinical scenarios. Due to the novel nature of the fragmentomic methods, their application has primarily focused on improving cancer detection. Here we demonstrate that using FrEIA in a cohort of 110 patients with resectable EAC monitored pre- and post-surgery, FrEIA has prognostic value at baseline for survival. When analyzing the 31 samples collected post-surgery, we demonstrated that 70% of patients with a FrEIA score above the detection threshold (13 out of 18 patients) developed a recurrence. Our results are comparable to mutation-based detection of tumor-derived cfDNA in resectable EAC *(7, 8)*. In a recent publication a ctDNA panel consisting of 77 genes was tested in 97 EAC patients. After filtering of CHIP variants, the panel showed high prognostic potential for disease-free survival (HR 5.35, 95% CI 2.10-13.63; P ≤ 0.0001) based on the post-surgery samples *(7)*. Another paper used an EAC tumor guided sequencing approach and found ctDNA status (positive vs. negative) to be prognostic at baseline for disease-free survival (p=0.042) *(8)*. In contrast to these two approaches, FrEIA metric does not require a buffy coat or a tumor biopsy, and has the potential to be easily implementable in the clinic and costs a fraction of tumor-informed sequencing. However, we must emphasize that the sensitivity of FrEIA is below those of tumor-informed sequencing methods (and bespoke sequencing panels that can reach parts per million fragments) *(28, 29)*. Furthermore, due to the nature of hybrid-capture sequencing, a combination of mutation analysis with FrEIA is possible and could result in improved tumor signal detection. In addition, FrEIA could assist in the detection, enrichment and characterization of bacterial, viral or pathogenic circulating DNA and mitochondrial DNA *(39)*. These DNA fragments are not associated with nucleosomes and are often highly fragmented below 100 bp *(33)*, while their FrESs remain to be explored. The microbiome diversity derived from circulating DNA can be linked to treatment efficiency in immuno-oncology, showing a promising diagnostic potential *(40)*.

Our results highlight that a cfDNA fragmentomic approach based on FrEIA has the potential to deliver sensitive detection of tumor-derived cfDNA using cheap low coverage WGS, although further validation in a larger cohort is needed. FrEIA can inform on the early detection of cancer and could contribute to addressing the unmet need of residual disease therapy decision making.

## MATERIAL AND METHODS

### Study design

A total of 572 plasma samples from 319 patients were retrieved across 18 cancer types, together with samples of 76 healthy controls and 10 plasma samples from patients with lung nodules or other lesions (**Table S1** and **S2**). Lung cancer patients and healthy individuals were recruited following informed consent via the Liquid Biopsy Center at the Amsterdam UMC, location VUmc and location AMC (METC U2019_035). Esophageal adenocarcinoma patients were recruited as part of the PERFECT trial or the BIOES esophageal and gastric cancer biobank (nCRT cohort) *(27)*. The PERFECT trial (METC 2016_325) and the BIOES biobank (METC 2013_241) have both received local approval from the medical ethical committee, resp. biobanking committee of the Academic Medical Center. Additional data were retrieved from a public database (EGA accession number: EGAS00001003258).

### Blood processing and DNA extraction

Blood samples were collected into EDTA-containing tubes and processed by a double-centrifugation protocol (1600 g for 10 minutes; 16000 g for 10 minutes) before storage at - 80°C. Blood samples collected locally in Amsterdam in EDTA coated tubes were processed using a double-centrifugation protocol (900 g for 15 minutes; 2500 g for 10 minutes). Supernatant plasma was carefully aliquoted in 0.5mL Nunc tubes before being stored at - 80°C.Plasma cfDNA was extracted using either the QIAamp Circulating Nucleic Acid Kit (QIAGEN; silica column-based) in cohort E or QIAsymphony DSP Circulating Nucleic Acid Kit (QIAGEN) for cohort L.

### Library preparation and sequencing

Plasma cfDNA was quantified using the cell-free DNA screentape kit and a Tapestation 4200 system (Agilent) or a BioAnalyzer HS chip and system (Agilent). Indexed sequencing libraries were prepared using 1-10 ng of DNA and the ThruPLEX-Plasma Seq kit or ThruPLEX-Tag Seq kit (Takara). Libraries were pooled in equimolar amounts and sequenced to <1x depth of coverage on a NovaSeq 6000 (Illumina) generating 150-bp paired-end reads from a S4 flowcell.

### Fragment End Analysis

Sequencing data were processed using a pipeline controlled by Snakemake (v. 5.14.0), and fragment ends were analysed using the FrEIA toolkit developed in our group [https://github.com/mouliere-lab/FrEIA.git]. In brief, adapters and indexes were trimmed using the bbduk.sh (v. 38.79) [https://sourceforge.net/projects/bbmap/] in paired mode with the ‘ktrim=r k=23 mink=11 hdist=1’ parameters and the adapter reference dataset provided with the software. The trimmed reads were mapped to the GRCh38 human genome assembly (GeneBank accession: GCA_000001405.28) using the bwa-mem (v. 0.7.17) [https://github.com/lh3/bwa]. Reads with a mapping quality lower than 5, unmapped reads, secondary mappings, chimeric and PCR duplicates were filtered with samtools (v. 1.12) [https://github.com/samtools/samtools]. Reads passing the filtration step were submitted for our custom pysam implementation, extracting the first 10 mapped bases from both ends of the remaining paired reads, together with the insert length, 5’ mapping position and contig. Fragments were categorized based on their first mapped nucleotide, the mapped 5’ trinucleotide sequence, as well as the first and last mapped nucleotides (**Figure 1A**). Fractions of these fragment categories were calculated for every sample.

### Fragment end motif entropy analysis

The FrES entropy was calculated for every sample as the normalized Shannon entropy *(26)* using the formula:

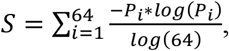

or as the Gini index using the formula:

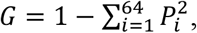

where *P*_*i*_ is the frequency of a specific trinucleotide ending.

### The FrEIA score calculation

Based on the observation that cfDNA fragment endings are non-random, and that cancer patients show a shift in fragment end sequences, we developed a single quantitative metric with the following formula:

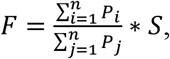

where *P*_*i*_ is the proportion of a given trinucleotide with significantly increased proportions in cancer, while *P*_*j*_ is the proportion of a given trinucleotide with significantly decreased proportions in cancer, while S is the normalized Shannon-entropy of the given sample. To select the trinucleotides with a significant increase or decrease in cancer we first selected samples with a SCNA-TF higher than 10% to ensure the tumor signal to noise ratio is high, and used these samples to calculate the log_10_ fold change of each trinucleotide proportion with the following formula:

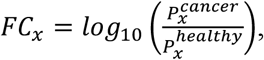

where *P*_*x*_ is the proportion of a given trinucleotide. Following this, we compared the mean proportion of each trinucleotide of the cancer cohort to the mean proportion of the same trinucleotide of the healthy cohort using the Wilcoxon Rank Sum Test and selected those that passed the 0.01 significance threshold. Those that had a FC lower than the 25% percentile were considered “significantly decreased in cancer”, while those that had a FC higher than the 75% percentile were considered “significantly increased in cancer”.

### Classification and predictive model

For the classification of baseline cancer samples from control samples we used the centralized log transformed proportions of mono- and trinucleotide sequence endings, together with the mononucleotide sequences on both ends and the normalized Shannon-entropy, Gini index of the trinucleotide endings and the FrEIA score. Superfluous features were eliminated by recursive feature elimination with cross-validation using the RFECV algorithm of Scikit-learn (v. 0.24.1) using the accuracy for scoring. To test the robustness of our model we performed both cross-validation with random sample selection on the whole dataset and independent validation, in brief:

#### Cross-validation with random sampling

Using Scikit-learn (v. 0.24.1) we split the data into 80% training-validation and 20% testing sets - stratified by the ‘cancer’ and ‘control’ categories - and scaled it using the StandardScaler. Splitting was repeated ten times using true random seeds generated on random.org: 67, 98, 6, 14, 62, 12, 28, 13, 80, 79. The training set was used for selecting the best estimator and parameters for our model (aka. hyperparameter tuning). For this we used GridSearchCV with 10-fold cross validation with sample shuffle and surveyed the parameter landscape of the KNeighbours, LogisticRegression, SupportVectorClassifier, RandomForrestClassifier and MLPClassifier estimators. The model with the highest score was selected and used to classify the prediction dataset, namely the SupportVectorClassifier with the parameters: ‘C’: 100, ‘gamma’: 1, ‘kernel’: ‘linear’ for the FrES metrics.

#### Independent validation

Using Scikit-learn (v. 0.24.1) we designated the R dataset as training and the L dataset as testing set. The E dataset containing only cancer cases was excluded. After scaling the data, we trained the previously selected SupportVectorClassifier with the previous parameters. For the classification of cancer types, we used both pre- and post-treatment samples. We performed cross-validation with random sampling on the R dataset and excluded the E and the L batches to avoid bias caused by possible batch effects. For a graphical representation of the classification sequence see **Figure 5A** and **Figure S13**.

### Synthetic dilutions

To determine the detection limits of our classifiers, we created a synthetic dilution series from one control and one cancer sample with 10% SCNA-TF as donor datasets, resulting in ten samples with SCNA-TF of 0%, 0.1%, 0.25%, 0.5%, 0.75%, 1%, 2.5%, 5%, 7.5% and 10%. Concurrently, we randomly downsampled the fragment counts of the dilutions to 1000, 10,000, 50,000, 100,000, 500,000 and 1 million fragments. The resulting 540 *in silico* dilutions and 60 controls were classified using the previously described SVM model with independent validation, trained on all baseline cancer samples and controls except the two donor samples. For performance evaluation we calculated the positive predictive value (PPV) and the negative predictive value (NPV) using the following equations:

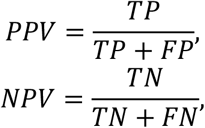

where *TP* is the number of true positives, *TN* is the number of true negative samples, *FP* is the count of false positives while *FN* is the number of false negatives.

### Statistical analysis and plotting

Fragment-end fractions per-definition represent compositional data, which lies in the simplex space, where standard statistical methods do not apply *(41)*. To free the data from this constraint we used centered log-ratio transformation (clr) from the ‘compositions’ package (v. 2.0.1) of R. For hypothesis testing on the clr-transformed data we used the two-sided Mann-Whitney U test with a significance level of 0.05, where not stated otherwise. When multiple hypotheses were tested, alpha values were adjusted using the Bonferroni method. Figures were plotted in RStudio (v. 1.3.1093) running R (v. 3.6.3) using ‘ggplot2’ (v. 3.3.3), ‘ggpubr’ (v. 0.4.0), ‘ggsci’ (v. 2.9) and ‘ggfortify’ (v. 0.4.11). The Kaplan-Meier analysis was performed using the R packages ‘survival’ (v 3.1-8) and visualized using ‘survminer’ (v. 0.4.9). Survival curves were calculated using the overall survival of the patients. The survival of patients who survived beyond the end of the study was censored.

## Data Availability

Datasets will be deposited in the European Genome-Phenome Archive (EGA) upon publication.

## SUPPLEMENTARY MATERIALS

Fig. S1: FrESs altered in cancer.

Fig. S2: the FrEIA score correlates with tumor fraction by cancer type

Fig. S3: the FrEIA score can detect early-stage cancer.

Fig. S4: the FrEIA score of patients with or without progression from the E data set.

Fig. S5: Kaplan-Meier progression curves and risk assessment of E samples with FrEIA score below and above the detection threshold measured at follow-up.

Fig. S6: Kaplan-Meier survival curves of patients with a FrEIA score above the detection threshold, patients with progression or patients with non-complete pathological response of E samples measured pre-treatment.

Fig. S7: the FrES metrics selected for the SVM classification.

Fig. S8: the performance of classification of cancer samples from control samples. Fig. S9: the R sample batch visualized using t-SNE.

Fig. S10: the FrES metrics selected for the SVM classification of one cancer type from all the others, but not controls in the R data set.

Fig. S11: ROC curves and AUROC values for cancer type classification using the FrEIA score or the FrES metrics.

Fig. S12: performance of the SVM model at different levels of TF and fragment count.

Fig. S13: classification of cancer stages, samples from patients with lung nodules and control patients.

Fig. S13: the workflow used to perform independent validation for classifying cancer from control.

Table S1: sample types and counts.

Table S2: the change in 5’ mononucleotide FrES proportions in cancer compared to control.

Table S3: the change in 5’ - 3’ mononucleotide FrES proportions in cancer compared to control.

Table S4: the change in 5’ trinucleotide FrES proportions in cancer compared to control.

Table S5: Spearman correlation coefficients of trinucleotide proportions and somatic copy number alteration-based tumor fraction (SCNA-TF) of baseline cancer samples.

Table S6: Spearman correlation coefficients of trinucleotide proportions and of the proportion of fragments with a size between 20-150 bp (P20-150) of baseline cancer samples.

Table S7: Spearman correlation coefficients of trinucleotide proportions and somatic copy number alteration-based tumor fraction (SCNA-TF) of cancer types with more than 10 samples at baseline.

Table S8: Spearman correlation coefficients of trinucleotide proportions and of the proportion of fragments with a size between 20-150 bp (P20-150) of cancer types with more than 10 samples at baseline.

## ADDITIONAL INFORMATIONS

## Acknowledgments

The authors are thankful to Mai Tran, Dr. Wendy Onstenk and the Amsterdam UMC Liquid Biopsy Center for the logistical support and advice. The authors are also thankful to Ilias Houda, Rimsha Shaikh, Ezgi Ulas for their help in annotating clinical information. Y.P., and F.M. are funded by the Amsterdam UMC Liquid Biopsy Center, an initiative made possible through the Stichting Cancer Center Amsterdam. The authors would like to thank Dr. Dineika Chandrananda for comments and discussions to improve the analysis of the ichorCNA algorithm. The authors would like to thank Dr. Caitrin Crudden for comments and discussions. This work was carried out on the Dutch national e-infrastructure with the support of SURF Cooperative.

## Funding

N.M. and F.M. are supported by a Dutch Cancer Fund (KWF-12822). The PERFECT study was financially supported by Hoffmann-La Roche Ltd., Basel, Switzerland. Analysis of cfDNA of the neoadjuvant chemoradiotherapy (nCRT) cohort was made possible through a grant of the Maag Lever Darm Stichting (SK18-32). Funders have no role in the design of the study.

## Author contributions

Conception and design: NM, FM

Experiments and data collection: YP, JR, SV, TV

Data processing: NM, YP, DB, FM

Software development: NM

Data analysis: NM, FM

Sample acquisition: TE, AC, MF, HL, IB

Funding acquisition: DMP, HL, IB, FM

Manuscript draft: NM, YP, FM

Manuscript revisions and comments: NM, YP, TE, DB, SV, JR, AC, TV, MF, DMP, HL, IB, FM

Supervision: FM

## Competing interests

F.M. is co-inventor on multiple patents related to cfDNA fragmentation analysis. Other co-authors have no relevant conflict of interests.

## Data and materials availability

Datasets will be deposited in the European Genome-Phenome Archive (EGA) upon publication. The FrEIA toolkit will be available on github [https://github.com/mouliere-lab/FrEIA.git].

## Notes

### Competing Interest Statement

Florent Mouliere is co-inventor on multiple patents related to cfDNA fragmentation analysis. Other co-authors have no relevant conflict of interests.

### Author Declarations

Amsterdam UMC ethics board

