## Supplementary Figures for "Genome-wide cell-free DNA termini in patients with cancer"

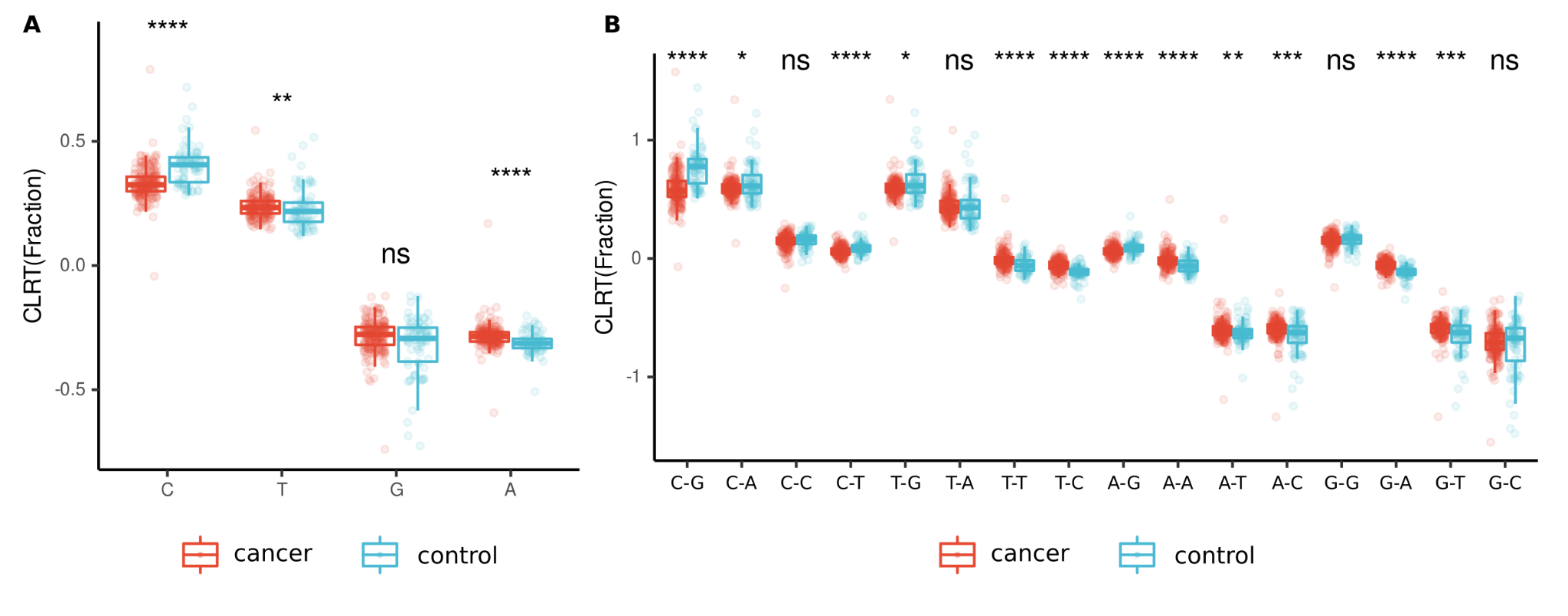

**Figure S1.** **FrESs altered in cancer.** **A.** 5’ mononucleotide FrES fractions in baseline cancer and control samples. **B.** The fraction of combined FrESs in baseline cancer and control samples. The first letter of the labels on the X axis denotes the 5’ end nucleotide, while the second letter the 3’ end nucleotide, both on the Watson strand. Fractions were transformed using the centered logration transformation method. P-values: ns: not significant, *: p < 0.05, **: p < 0.01, ***: p < 0.005, ****: p < 0.001.

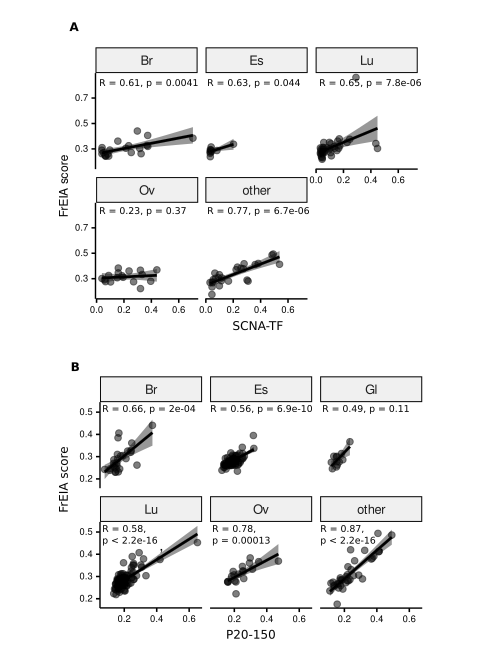

**Figure S2.** **The FrEIA score correlates with tumor fraction by cancer type.** The tumor fraction of baseline cancer samples was measured either by **A.** somatic copy number analysis (SCNA-TF) or **B.** the proportion of fragments between 20 and 150 bp (P20-150). For SCNA-TF samples with a TF > 3% are shown. Cancer types: Br: breast cancer, Es: esophageal cancer, Gl: glioblastoma, Lu: lung cancer, Ov: ovarian cancer. Cancer types with less than 10 samples at baseline were merged into the ‘other’ category.

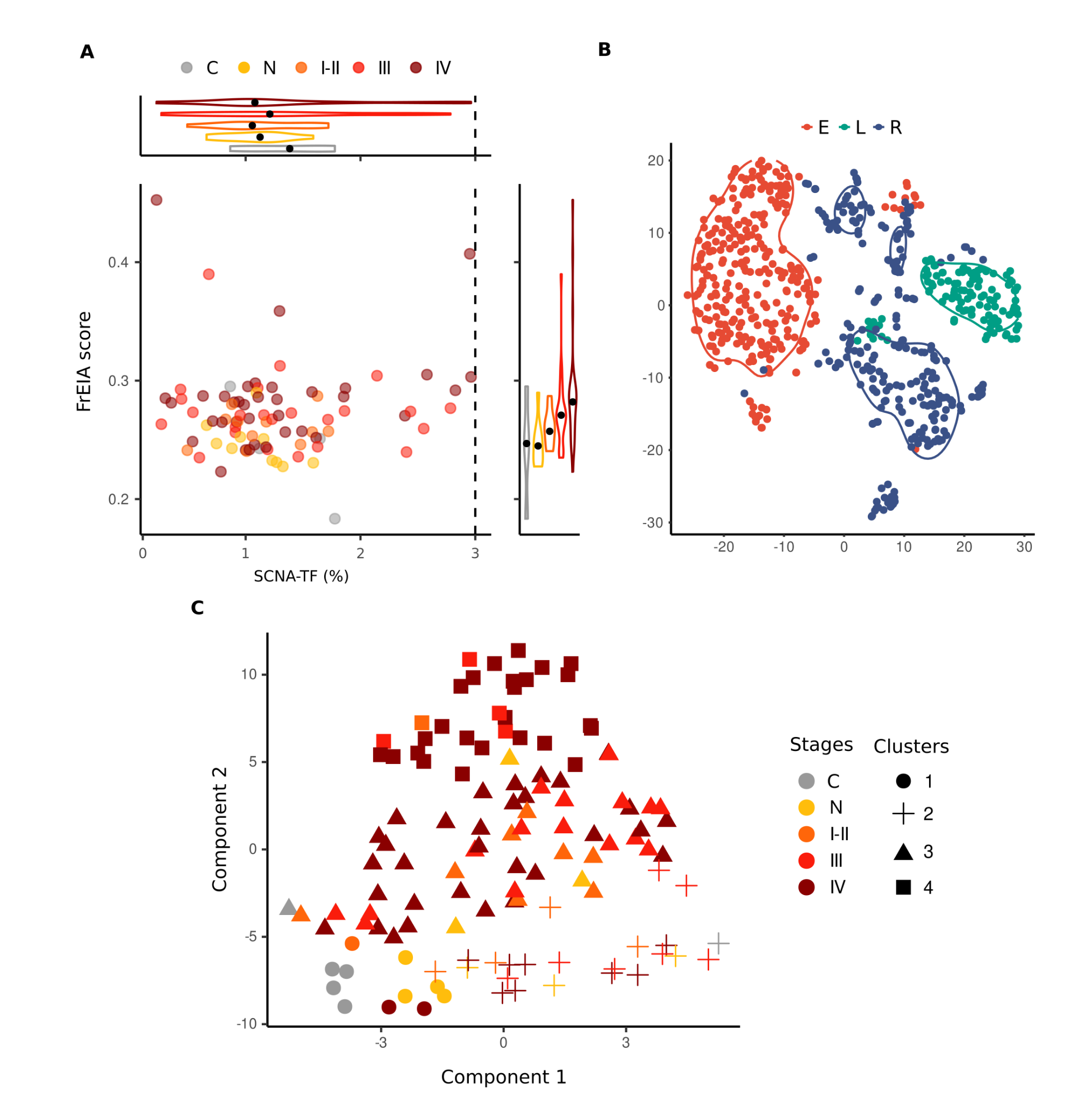

**Figure S3. The FrEIA score can detect early stage cancer.** **A.** The FrEIA score separates lung cancer stages with an SCNA-TF below the detection threshold 3%. The detection threshold is represented by the vertical dashed line. **B.** Sample batches visualized using t-SNE on the FrES metrics of the three datasets used in this study. E: esophageal cancer dataset, L: lung cancer dataset, R: retrieved dataset. **C.** Classification of cancer stages, samples from patients with lung nodules and control patients. Samples were grouped in 4 clusters using the k-means method and visualized using t-SNE. Cancer stages: C: control (n = 6), N: nodule or lesion (n = 10), stages I-II: n = 14, stage III: n = 27, stage IV: n = 66.

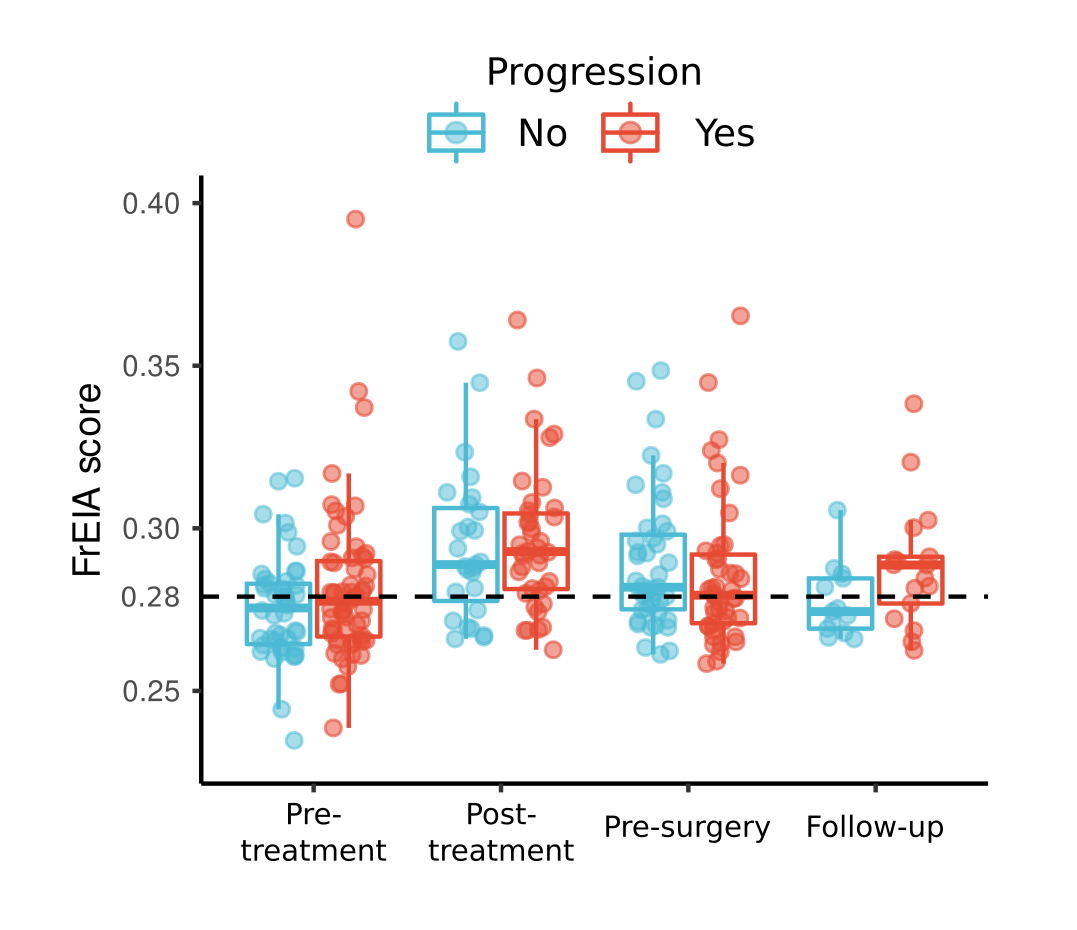

**Figure S4. The FrEIA score of patients with or without progression from the E data set.** The vertical dashed line represents a conservative threshold for FrEIA score derived from in-silico dilutions.

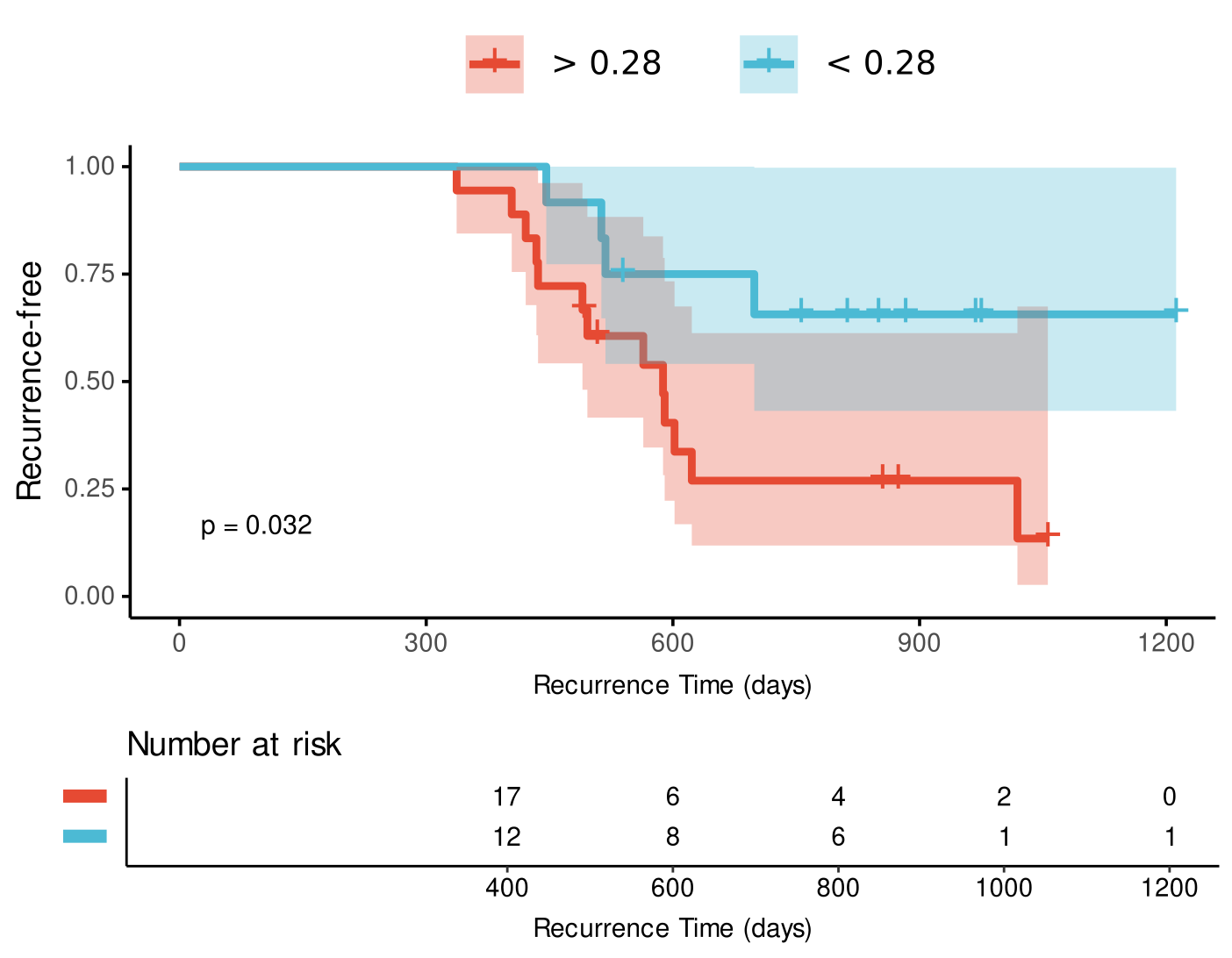

**Figure S5. Kaplan-Meier progression curves and risk assessment of E samples with FrEIA score below and above the detection threshold measured at follow-up.**

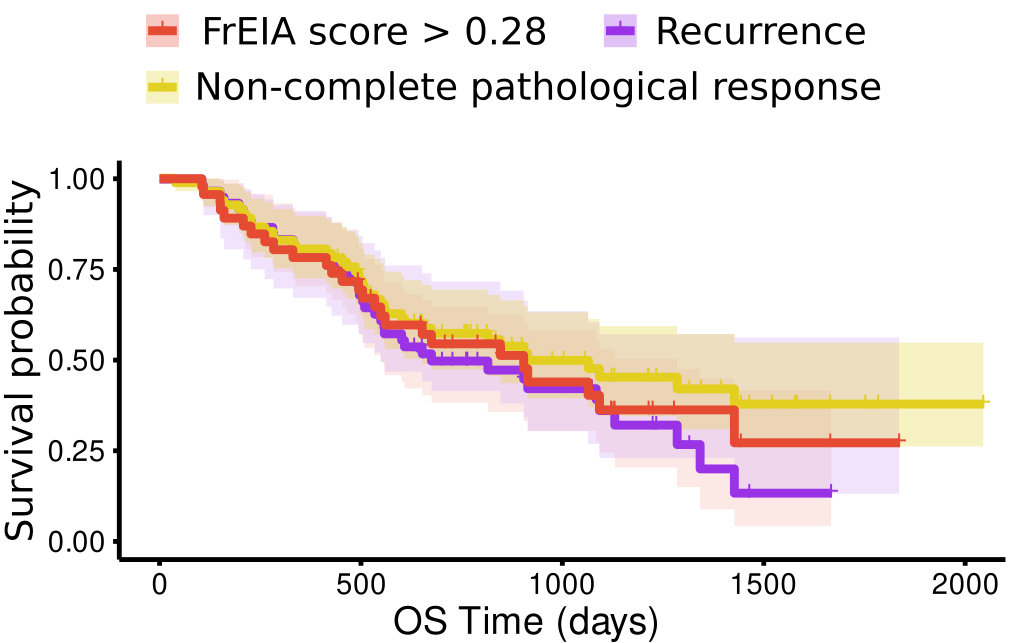

**Figure S6. Kaplan-Meier survival curves of patients with a FrEIA score above the detection threshold, patients with progression or patients with non-complete pathological response of E samples measured pre-treatment.**

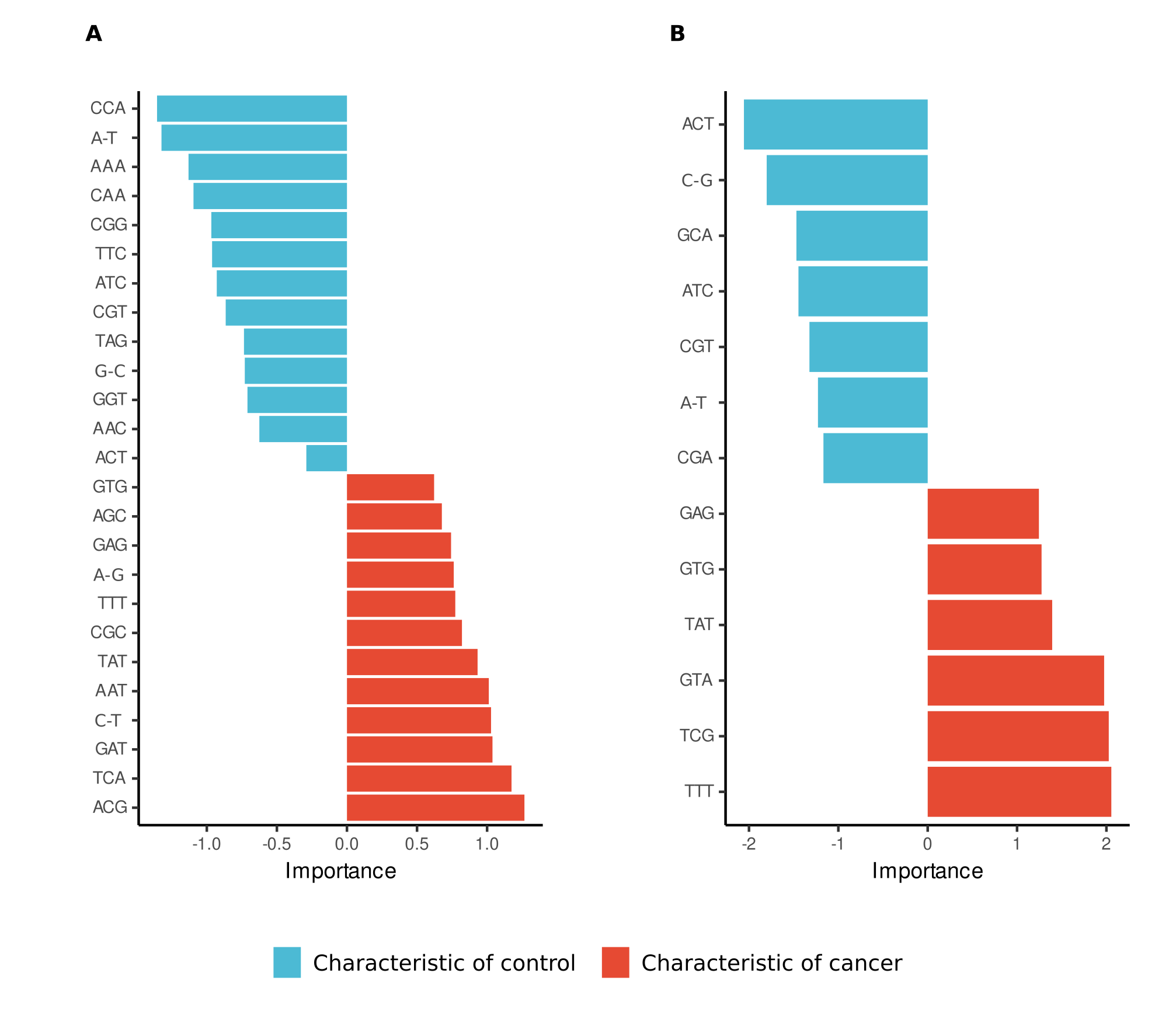

**Figure S7. The FrES metrics selected for the SVM classification. A.** The FrES metrics selected for the cross-validation-based approach. **B.** The FrES metrics selected for the independent validation-based approach. Feature selection was performed by recursive feature elimination (RFE). The importance is the SVM coefficient calculated for the specific feature.

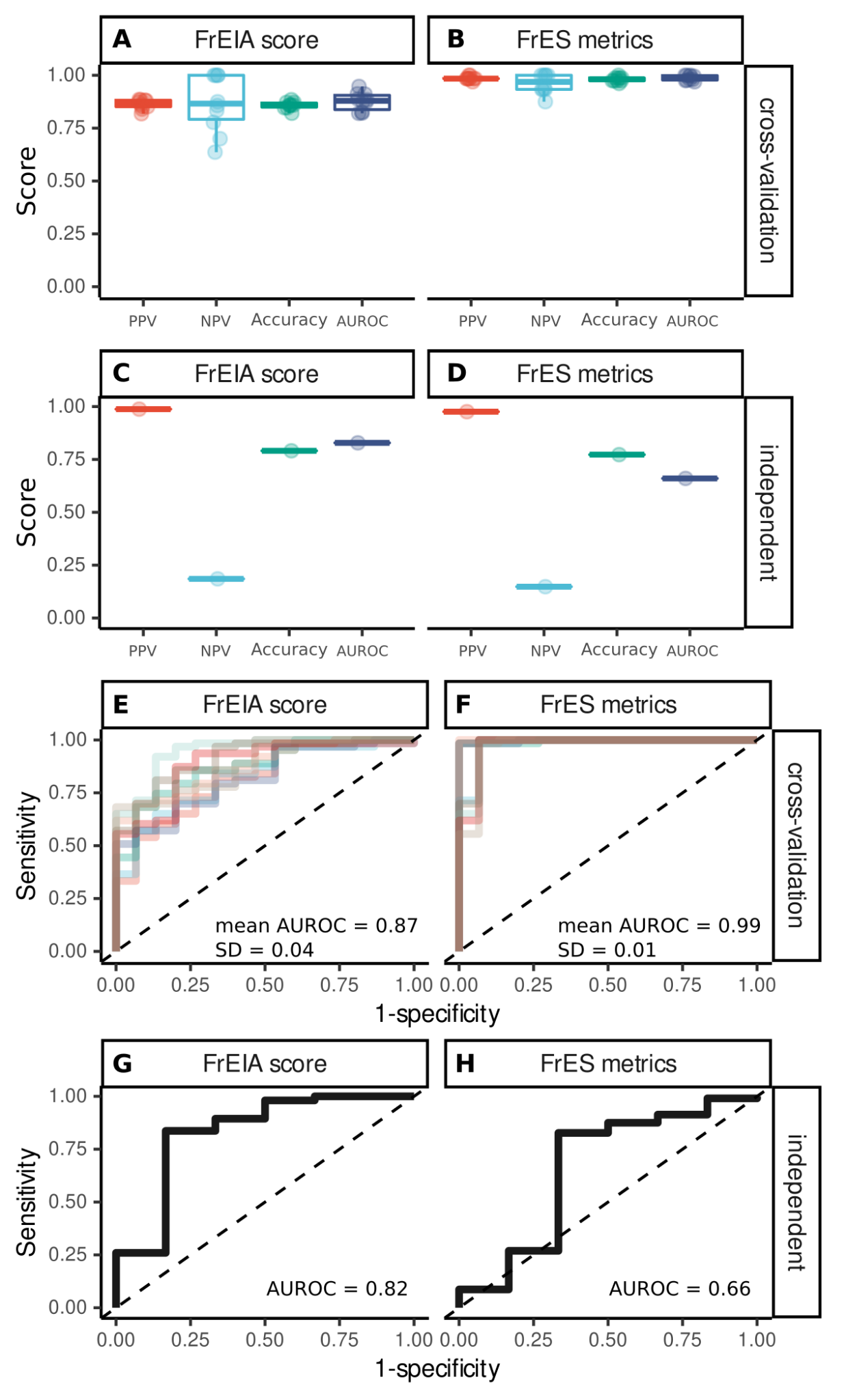

**Figure S8. The performance of classification of cancer samples from control samples.** SVM classifiers were trained using 10 fold cross-validation with random sampling using all pre-treatment samples (panels **A**, **B** and **E**, **F**) and independent validation using the R samples for training and the L dataset for predictions (panels **C**, **D** and **G,** **H**). Models were created using the FrEIA score (panels **A**, **C**, **E** and **G**) or the FrES metrics (panels **B**, **D**, **F** and **H**).

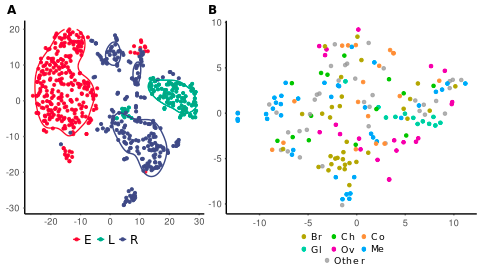

**Figure S9. The R sample batch visualized using t-SNE.** FrES metrics and FrEIA score-based visualization of the R dataset. Cancer types are shown: Br: breast, Ch: cholangeal, Co: colorectal, Gl: glioma, Ov: ovarian, Me: melanoma.

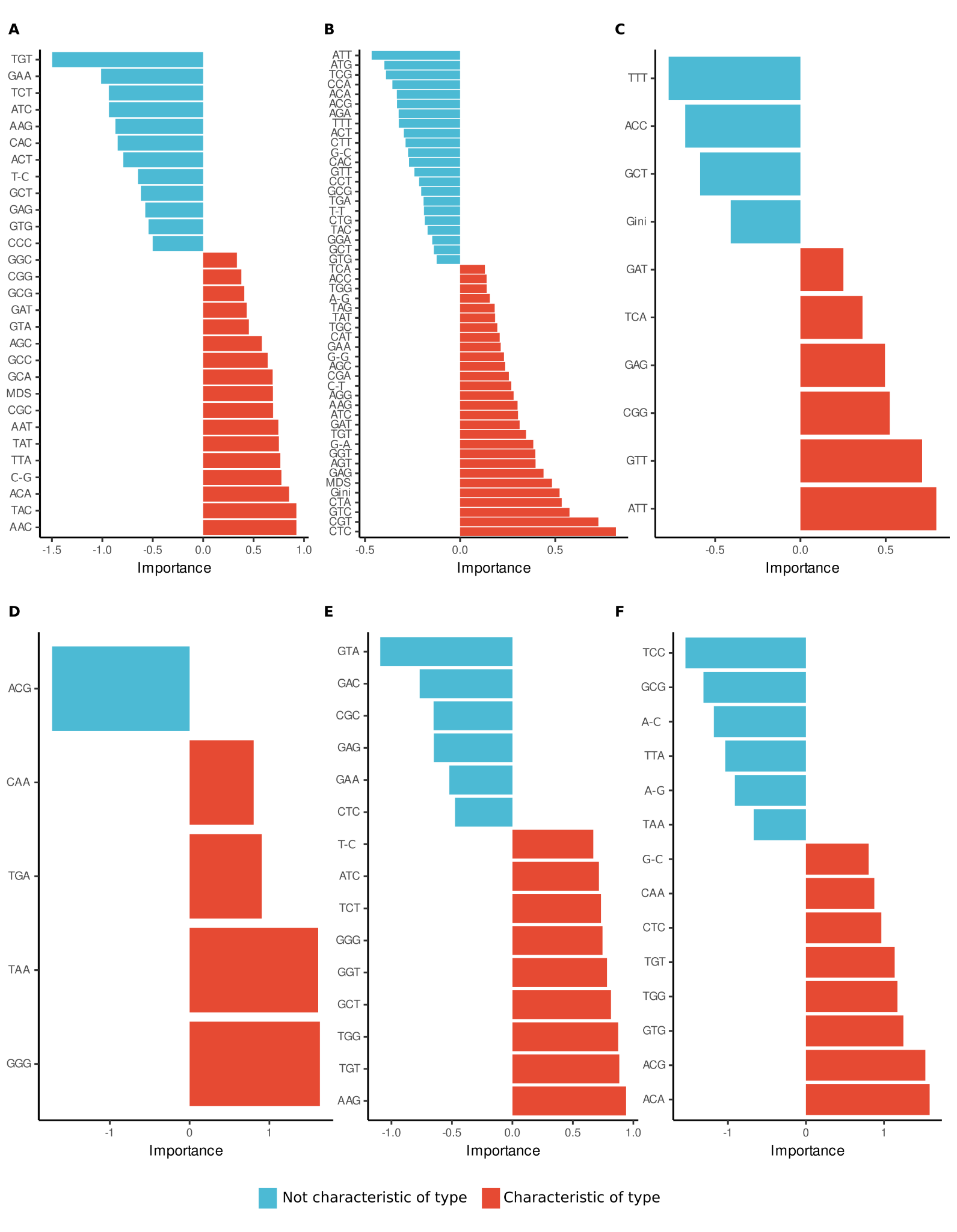

**Figure S10. The FrES metrics selected for the SVM classification of one cancer type from all the others, but not controls in the R data set. A.** breast, **B.** cholangiocarcinoma, **C.** colorectal, **D.** glioblastoma, **E.** melanoma, **F.** ovarian cancer. Feature selection was performed by recursive feature elimination (RFE). The importance is the SVM coefficient of the specific feature.

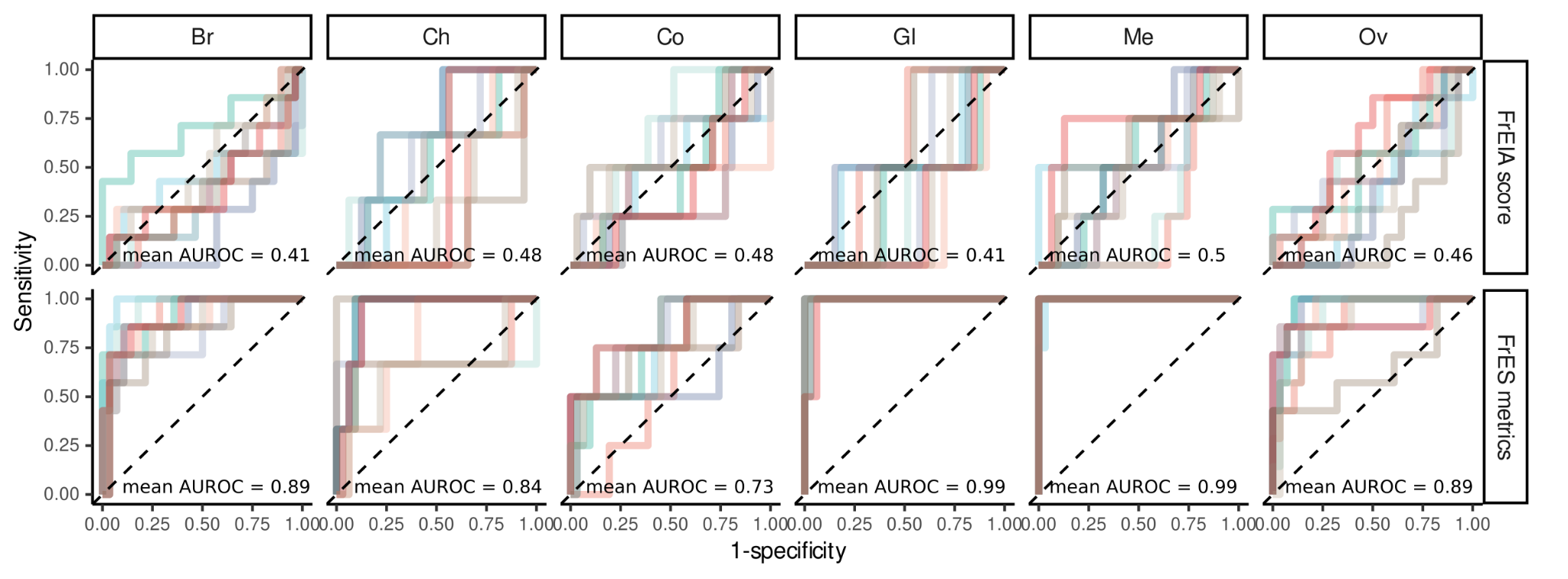

**Figure S11. ROC curves and AUROC values for cancer type classification using the FrEIA score or the FrES metrics.** Cancer types: Br: breast, Ch: cholangeal, Co: colorectal, Gl: glioma, Me: melanoma, Ov: ovarian.

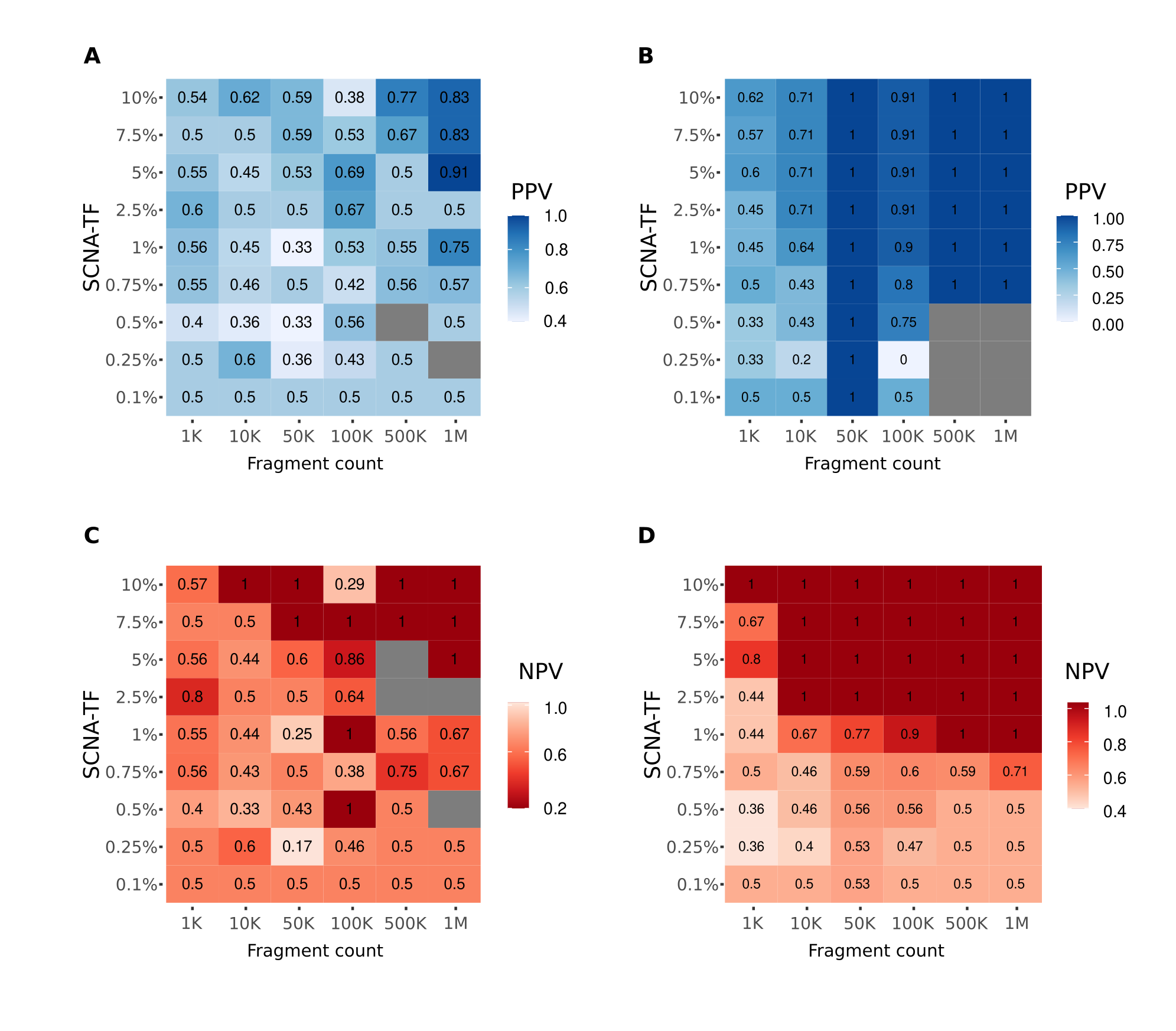

**Figure S12. Performance of the SVM model at different levels of TF and fragment count.** The positive predictive value (PPV) of the (**A.**) FrES metrics-based and (**B.**) FrEIA score-based models. The negative predictive value (NPV) of the (**C.**) FrES metrics-based and (**D.**) FrEIA score-based models. Gray cells hold values where the amount of true-positives and false-positives were 0 and thus a PPV, and where the amount of true-negatives and false-negatives were 0 and thus a NPV could not be computed.

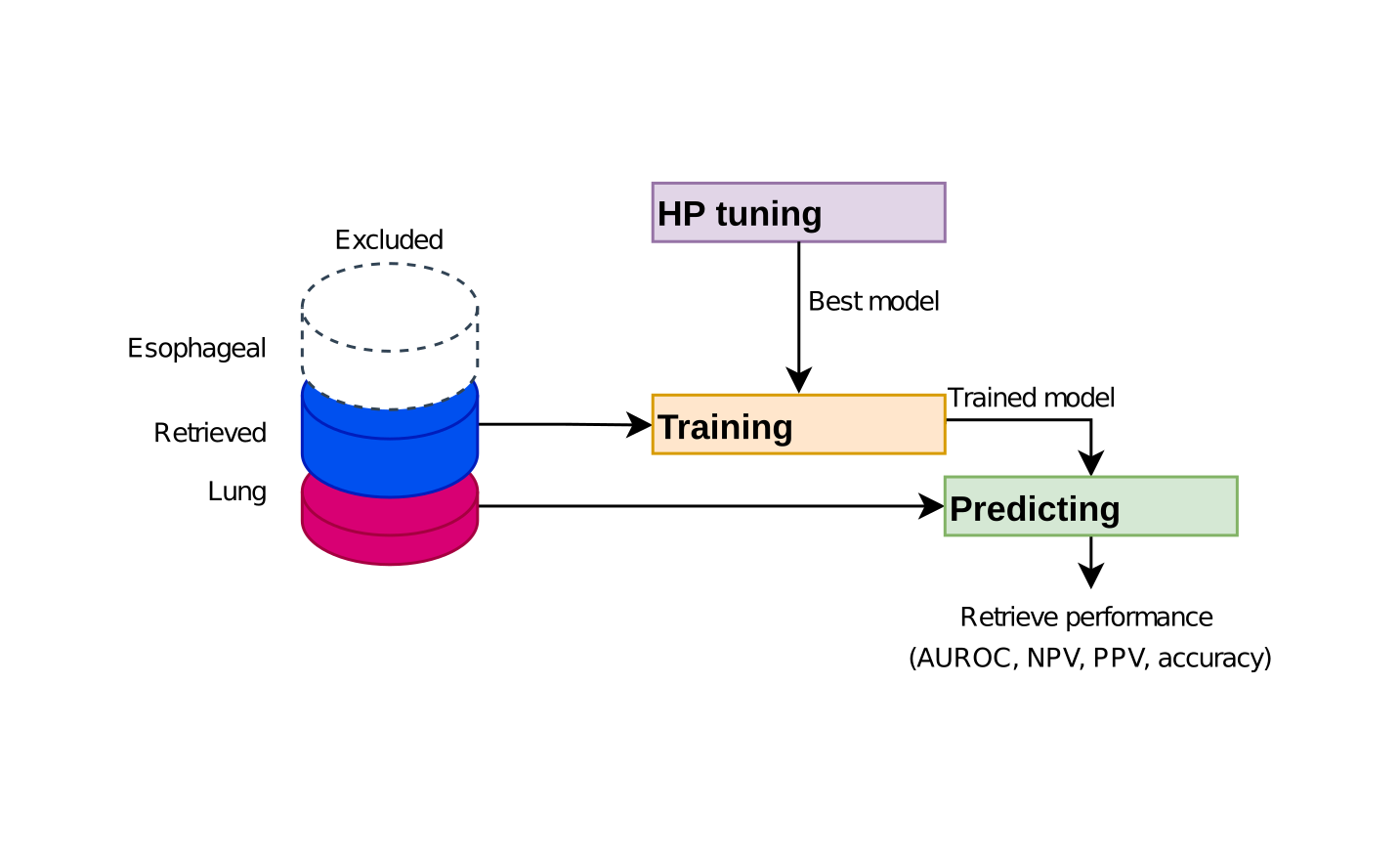

**Figure S13. The workflow used to perform** independent validation for classifying cancer from control. HP tuning: hyper-parameter tuning for model selection.

### Supplementary table legends

**Table S1. Sample types and counts.**

**Table S2. The change in 5’ mononucleotide FrES proportions in cancer compared to control.**

**Table S3. The change in 5’ - 3’ mononucleotide FrES proportions in cancer compared to control.**

**Table S4. The change in 5’ trinucleotide FrES proportions in cancer compared to control.**

**Table S5.** **Spearman correlation coefficients of trinucleotide proportions and somatic copy number alteration-based tumor fraction (SCNA-TF) of baseline cancer samples.**

**Table S6.** **Spearman correlation coefficients of trinucleotide proportions and of the proportion of fragments with a size between 20-150 bp (P20-150) of baseline cancer samples.**

**Table S7. Spearman correlation coefficients of trinucleotide proportions and somatic copy number alteration-based tumor fraction (SCNA-TF) of cancer types with more than 10 samples at baseline.**

**Table S8. Spearman correlation coefficients of trinucleotide proportions and of the proportion of fragments with a size between 20-150 bp (P20-150) of cancer types with more than 10 samples at baseline.**

### Supplementary methods

**Testing for batch effects**

To test for potential classification bias ^24^ resulting from deviations in cfDNA handling in the three sample batches, we analyzed our samples using the *t*-distributed stochastic neighbor embedding (t-SNE) method. This suggested that FrES metrics can be sensitive to preanalytical variations (**Figure S3B**). To further test if our datasets can be used for classification we performed both cross-validation with random sample selection on the whole baseline dataset (**Figure 3A**) and an independent validation using the R dataset as training and the L dataset as test set (see Methods and **Figure S13**). Our model performed poorly with independent validation when trained on the FrES metrics (**Figure S8**). Intriguingly, when using the FrEIA score solely, the performance is close to the performance of the cross-validated model, but the Negative Predictive Value (NPV) of the model is much lower (independent validation: AUROC = 0.83, NPV = 0.19; cross validation: mean AUROC = 0.88, SD = 0.04, NPV = 0.87) (**Figure S8C, D, G** and **H**). This confirms that FrES metrics (a total of 82 variables) can be strongly influenced by the preanalytical conditions, and classification between sample batches need to be done with precaution.
